# Spread of Gamma (P.1) sub-lineages carrying Spike mutations close to the furin cleavage site and deletions in the N-terminal domain drives ongoing transmission of SARS-CoV-2 in Amazonas, Brazil

**DOI:** 10.1101/2021.09.12.21263453

**Authors:** Felipe Gomes Naveca, Valdinete Nascimento, Victor Souza, André de Lima Corado, Fernanda Nascimento, George Silva, Matilde Mejía, Maria Júlia Brandão, Ágatha Costa, Débora Duarte, Karina Pessoa, Michele Jesus, Luciana Gonçalves, Cristiano Fernandes, Tirza Mattos, Ligia Abdalla, João Hugo Santos, Alex Martins, Fabiola Mendonça Chui, Fernando Fonseca Val, Gisely Cardoso de Melo, Mariana Xavier Simão, Vanderson de Souza Sampaio, Maria Paula Mourão, Marcus Vinícius Lacerda, Érika Lopes Rocha Batista, Alessandro Leonardo Álvares Magalhães, Nathânia Dábilla, Lucas Carlos Gomes Pereira, Fernando Vinhal, Fabio Miyajima, Fernando Braga Stehling Dias, Eduardo Ruback dos Santos, Danilo Coêlho, Matheus Ferraz, Roberto Lins, Gabriel Luz Wallau, Edson Delatorre, Tiago Gräf, Marilda Mendonça Siqueira, Paola Cristina Resende, Gonzalo Bello

## Abstract

The Amazonas was one of the most heavily affected Brazilian states by the COVID-19 epidemic. Despite a large number of infected people, particularly during the second wave associated with the spread of the Variant of Concern (VOC) Gamma (lineage P.1), SARS-CoV-2 continues to circulate in the Amazonas. To understand how SARS-CoV-2 persisted in a human population with a high immunity barrier, we generated 1,188 SARS-CoV-2 whole-genome sequences from individuals diagnosed in the Amazonas state from 1st January to 6th July 2021, of which 38 were vaccine breakthrough infections. Our study reveals a sharp increase in the relative prevalence of Gamma plus (P.1+) variants, designated as Pango Lineages P.1.3 to P.1.6, harboring two types of additional Spike changes: deletions in the N-terminal (NTD) domain (particularly **Δ**144 or **Δ**141-144) associated with resistance to anti-NTD neutralizing antibodies or mutations at the S1/S2 junction (N679K or P681H) that probably enhance the binding affinity to the furin cleavage site, as suggested by our molecular dynamics simulations. As lineages P.1.4 (S:N679K) and P.1.6 (S:P681H) expanded (Re > 1) from March to July 2021, the lineage P.1 declined (Re < 1) and the median Ct value of SARS-CoV-2 positive cases in Amazonas significantly decreases. Still, we found no overrepresentation of P.1+ variants among breakthrough cases of fully vaccinated patients (71%) in comparison to unvaccinated individuals (93%). This evidence supports that the ongoing endemic transmission of SARS-CoV-2 in the Amazonas is driven by the spread of new local Gamma/P.1 sub-lineages that are more transmissible, although not more efficient to evade vaccine-elicited immunity than the parental VOC. Finally, as SARS-CoV-2 continues to spread in human populations with a declining density of susceptible hosts, the risk of selecting new variants with higher infectivity are expected to increase.

## Introduction

The Amazonas was one of the most heavily affected Brazilian states by the COVID-19 epidemic, and by 31st July 2021, 13,531 deaths had been reported (1). The COVID-19 epidemic in Amazonas was characterized by two waves of exponential growth (**Figure 1 A**). The first one started in March 2020 and peaked around early May 2020, and was primarily associated with the introduction and dissemination of lineages B.1.195 and B.1.1.28 (2). The second one started in December 2020 and peaked around early February 2021 and was associated with the local emergence and rapid spread of the Gamma/P.1 lineage, a more transmissible SARS-CoV-2 Variant of Concern (VOC) firstly detected in Japanese travelers returning from the Amazonas State, Brazil (2-4). Since mid-February 2021, the number of SARS-CoV-2 deaths dropped and then remained roughly stable (7-day average <20) between May and July 2021.

**Figure 1.**
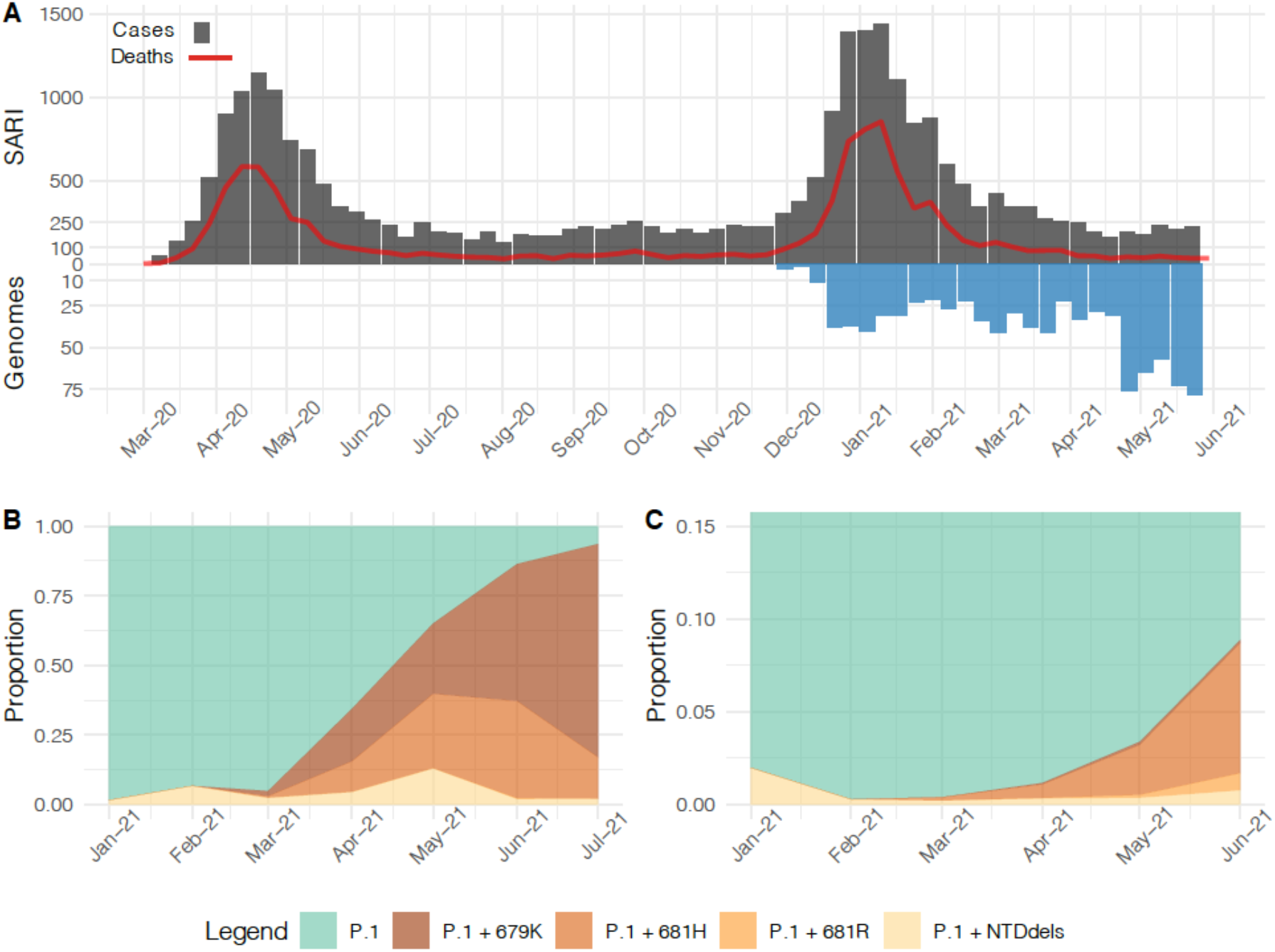
Temporal distribution and genetic diversity of SARS-CoV-2-positive samples from the Amazonas state during 2021. **A**, Graph depicting the temporal evolution of SARI cases and SARI deaths based on the date of symptom onset (source, http://info.gripe.fiocruz.br) as a proxy for the COVID-19 epidemic curve in Amazonas state, along with the number of SARS-CoV-2 whole-genome sequences generated between January and July 2021. **B**, Relative frequency of different P.1 lineage variants among SARS-CoV-2 positive cases sequenced in the Amazonas. **C**, Relative frequency of different P.1 lineage variants among SARS-CoV-2 P.1 Brazilian sequences detected outside the Amazonas states.

Despite many infected people in the two waves of the COVID-19 pandemic and the relatively high percentage of partial (32%) and fully (18%) vaccinated individuals in the Amazonas state on 31st July 2021 (5), the virus continues to circulate at a roughly steady-state level of ∼500 SARS-CoV-2 positive cases per day (7-day rolling average) from early May to mid-July, 2021 (6). This endemic pattern of virus transmission supports the assumption that population immunity acquired in Amazonas, either through vaccinations or natural SARS-CoV-2 infection, is sufficient to prevent new exponential growth but not to stop the spread of the virus. We hypothesize that the endemic transmission of the VOC Gamma/P.1 in the Amazonas state allowed the rise of second-generation variants presenting a higher herd-immunity threshold than the parental virus.

To test this hypothesis, we combined phylodynamic approaches with epidemiological data to track mutations accumulated in SARS-CoV-2 whole-genomes recovered from individuals living in the Amazonas state diagnosed between January and July 2021. We demonstrate that persistent SARS-CoV-2 circulation in Amazonas was associated with a sharp increase in the relative prevalence of local P.1 plus (P.1+) variants harboring deletions in the N-terminal domain (NTD) or mutations at the S1/S2 junction of the Spike protein, while the parental P.1 lineage has been through a progressive extinction process since May 2021. New P.1+ lineages are more transmissible than the parental one, but natural and vaccine derived immunity are contributing to limit the spread of these lineages in the Amazonas state.

## Results

### The ongoing evolution of lineage P.1 in the Amazonas state, Brazil

To better understand the recent evolution of VOC P.1 spreading in the Amazonas state, we sequenced the virus genome from 1,137 patients representing a random sample of SARS-CoV-2 positive cases diagnosed from 1st January to 6th July 2021. The Amazonas state health surveillance foundation sent SARS-CoV-2 positive samples from different municipalities for sequencing at FIOCRUZ Amazônia, part of the local health genomics network (REGESAM) and the consortium FIOCRUZ COVID-19 Genomics Surveillance Network of the Brazilian Ministry of Health (http://www.genomahcov.fiocruz.br/). Furthermore, we also sequenced the virus from 51 individuals living in Manaus who were part of a cohort of patients receiving the CoronaVac vaccine (CovacManaus https://www.ipccb.org/covacmanaus). This resulted in a total of 1,188 SARS-CoV-2 high-quality (<1% of undetermined “N” bases) whole-genome (>98% coverage) sequences, representing 0.6% of all laboratory-confirmed SARS-CoV-2 cases in the Amazonas state from 1st January to 6th July 2021 (*n* = 202,773) (**Figure 1 A**).

Our genomic survey confirms that VOC Gamma was the most prevalent lineage representing 99.7% (1,185 of 1,188 genomes) of all samples sequenced in the Amazonas state across the study period. We identified over 127 distinct amino acid (AA) substitutions and deletions in the S protein in addition to those that define lineage P.1, most of them present in less than ten samples (**Supplementary Table 1**). AA deletions covering in the NTD region (**Δ**144, **Δ**141-144, and **Δ**138-143) and substitutions at the S1/S2 junction (N679K, P681H, and P681R), however, were particularly prevalent and sharply increase from January to May 2021 (**Figure 1 B**). The P.1+NTDdel variants increased from 1.6% in January to 12.4% in May 2021, while the P.1+N679K and P.1+P681H variants increased from 0% to 25.4% and 17.6%, respectively, in the same period. During June-July 2021, the proportion of P.1+N679K genomes continued to increase up to 76.9%, the relative frequency of P.1+P681H genomes increased up to 37.1% in June then decrease to 13.5% in July, while the frequency of variants P.1+NTDdel dropped to 2.1%. Inspection of P.1 sequences available at EpiCoV database in the GISAID (https://www.gisaid.org/ 4) on July 22nd, 2021, also detected an increased frequency of variants P.1+NTDdel, P.1+N679K, P.1+P681H/R in other Brazilian states, but in a much lower prevalence than in the Amazonas (**Figure 1 C**).

**Table 1.** P.1+ lineage-defining mutations present in >95% of sequences.

| P.1 sub-lineage | First detected | Nucleotide | Amino acid |
| --- | --- | --- | --- |
| P.1+Δ144<sub>AM</sub> | 28th Apr 2021 | C12513T<br>Δ21992-21994 | ORF1a:T4083M<br>S:Δ141-144 |
| P.1.3 | 12th Mar 2021 | C5526T<br>C16193T<br>A21979T<br>Δ21983-21994<br>T27826C<br>C27942T | ORF1a:T1754I<br>ORF1b:P909L<br>-<br>S:Δ141-144<br>ORF7b:M24T<br>ORF8:H17Y |
| P.1.4 | 19th Mar 2021 | T23599G | S:N679K |
| P.1.5 | 3rd Apr 2021 | C3117T<br>A18945G<br>T23599A | ORF1a:T951I<br>-<br>S:N679K |
| P.1.6 | 26th Mar 2021 | C10615T<br>C15714T<br>C23604A | -<br>-<br>S:P681H |
| P.1.7 | 12th Mar 2021 | C1912T<br>C16293T<br>C23604A | -<br>-<br>S:P681H |
| P.1.8 | 24th May 2021 | T592C<br>A4040G<br>C9891T<br>A20055G<br>C22971A<br>C23604G<br>G25266T<br>C26039T<br>G27915T | -<br>ORF1a:I1259V<br>ORF1a:A3209V<br>-<br>S:T470N<br>S:P681R<br>S:C1235F<br>ORF3a:S261L<br>ORF8:G8stop |

### Identification of major P.1+ lineages in Amazonas and other Brazilian states

To determine if the most frequent P.1 mutations detected in Brazil resulted from independent convergent mutations events, we combined the Amazonian P.1 sequences generated in this and previous studies with P.1+NTDdel (**Δ**144, **Δ**143-144, and **Δ**141-144), P.1+N679K and P.1+P681H/R sequences detected in other Brazilian states that were available at the EpiCoV database in GISAID (https://www.gisaid.org/). The Maximum Likelihood (ML) phylogenetic analysis supports that NTD deletions around position Y144 arose multiple (n > 20) times during the evolution of lineage P.1 in Brazil, in agreement with our previous observations (7), as well as mutations S:N679K (n > 5), S:P681H (n > 4) and S:P681R (n > 3) (**Figure 2**). This analysis further revealed four well-supported (aLRT = 76-99%) monophyletic P.1+ sub-clades in Amazonas; some of which received the Pango Lineage designation P.1.3 to P.1.6 (**Figure 2**). Lineage P.1.3 comprises most P.1+**Δ**141-144 sequences from the Amazonas state (n = 29/33, 88%). Lineage P.1.4 comprises most P.1+N679K sequences from the Amazonas state (n = 187/197, 95%) and three P.1+N679K sequences from Rio de Janeiro. Lineage P.1.5 comprises the remaining P.1+N679K sequences from the Amazonas state (n = 10/197, 5%) and two P.1+N679K sequences from Roraima and São Paulo states. Lineage P.1.6 comprises most P.1+P681H sequences from the Amazonas state (n = 208/209, 99%) and one P.1+P681H sequence from Rio de Janeiro. The fourth lineage, designated as P.1+**Δ**144_AM_, comprises about half of P.1+**Δ**144 sequences from Amazonas (n = 12/29, 41%). Our analyses also revealed two major well-supported (aLRT = 76-99%) P.1+ lineages designated as P.1.7 and P.1.8 that comprises most P.1+P681H (n = 227/234, 97%) and P.1+P681R (n = 13/20, 65%) sequences detected outside the Amazonas state, respectively. Analysis of the mutational profile reveals a variable number of lineage-defining mutations ranging from one in P.1.4 to nine in P.1.8 (**Table 1**). Interestingly, lineages P.1.4 and P.1.5 displayed the same lineage-defining AA substitution (N679K) but different nucleotide mutations (T23599G and T23599A). Most P.1+ sub-clades displayed only one AA lineage-defining mutation in the S protein, except P.1.8 that displayed three mutations (T470N, P681R, and C1235F).

**Figure 2.**
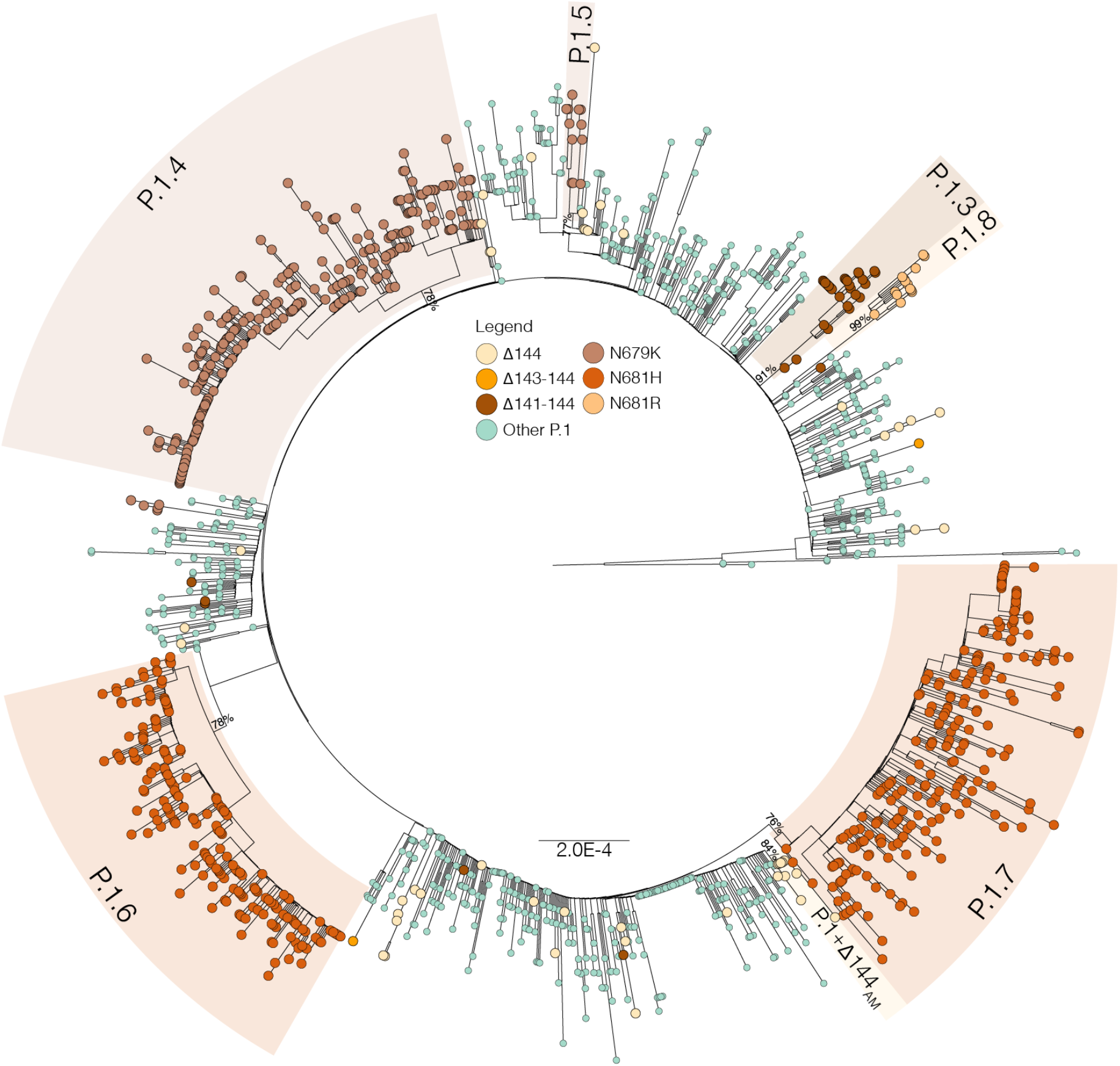
Maximum likelihood phylogenetic tree of P.1 Amazonian sequences and P.1+NTDdel, P.1+N679K, and P.1+P681H sequences detected outside Amazonas. Tips were colored according to the S mutations as indicated in the legend. Major P.1 sub-lineages carrying additional mutations/deletions in the S protein were highlighted with colored boxes. The aLRT support values are indicated in key branches, and branch lengths are drawn to scale with the bar indicating nucleotide substitutions per site.

### Onset date and geographic spread of major Brazilian P.1+ lineages

The analysis of temporal structure revealed that the overall divergence of the new P.1+ lineages is comparable with the divergence of other contemporaneous P.1 genomes circulating in the Amazonas, thus supporting an homogenous evolutionary rate among lineages (**Figure 3 A**). Bayesian time-scaled reconstruction estimated the emergence of the lineages P.1.3 at 7th Mar 2021 (95% HPD: 21th Feb – 12th Mar 2021), P.1.4 at 13th Feb 2021 (95% HPD: 9th Jan – 10th Mar 2021), P.1.5 at 15th Mar 2021 (95% HPD: 21th Feb – 30th Mar 2021), P.1.6 at 3rd March 2021 (95% HPD: 3rd Feb – 24th Mar 2021), P.1.7 on 16th Feb 2021 (95% HPD: 19th Jan – 5th Mar 2021), P.1.8 at 11th May (95% HPD: 1st May – 19th May 2021), and P.1+**Δ**144_AM_ at 21st Apr 2021 (95% HPD: 6th Apr – 28th Apr 2021) (**Figures 3 B – 3 E**). Of note, the estimated median tMRCA of most P.1+ lineages circulating in Amazonas (13th Feb - 15th Mar) coincided with the declining phase of the second COVID-19 epidemic wave in the state.

**Figure 3.**
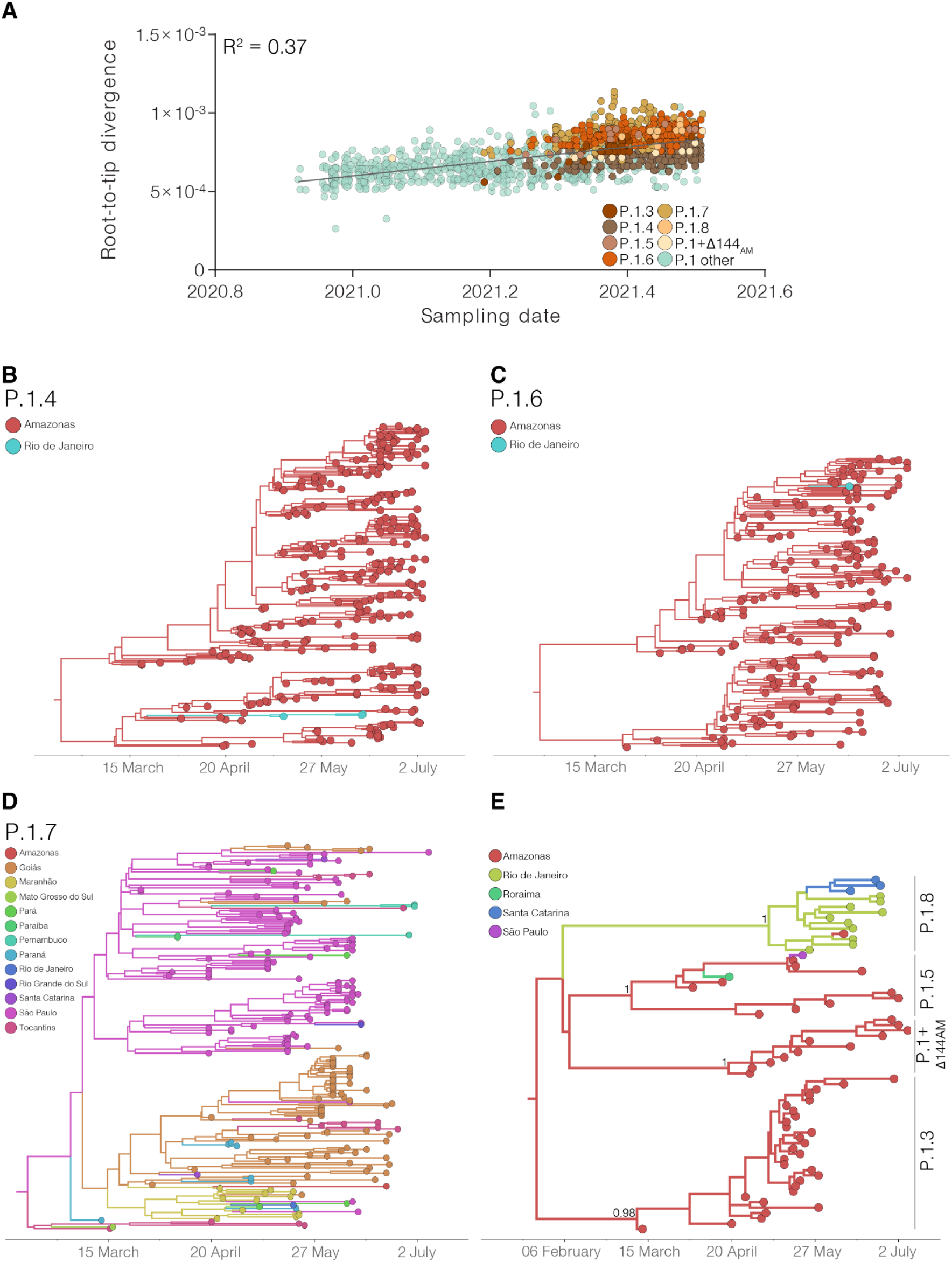
Temporal structure and phylogeographic reconstruction of the P.1+NTDdel, P.1+N679K, and P.1+P681H clades. **A**, root-to-tip regression of genetic divergence against dates of sample collection. P.1 sequences were colored green, while each P.1 subclade carrying deletions or additional mutations in S protein was colored following the legend. Time-resolved maximum clade credibility phylogenies of each P.1 subclade defined in the ML analysis: **B**, P.1.4; **C**, P.1.6; **D**, P.1.7; **E**, minor clades P.1.3, P.1.5, P.1.8 and P.1+**Δ**141-144_AM_. Tips and branches colors indicate the sampling state and the most probable inferred state of the nodes, respectively, as indicated in the legend for each tree. Bayesian posterior probabilities are indicated in key branches. All horizontal branch lengths are time-scaled, and the tree was automatically rooted under the assumption of the molecular clock model.

As expected, phylogeographic reconstructions traced the Amazonas state as the most probable source location (PSP = 1) of lineages P.1.3, P.1.4, P.1.5, P.1.6, and P.1 +**Δ**144_AM_. These P.1 sub-lineages remained mostly restricted to the Amazonas state, except for a few sporadic disseminations to the Rio de Janeiro (lineages P.1.4 and P.1.6), Roraima (lineage P.1.5) and São Paulo (lineage P.1.5) states (**Figures 3 B, 3 C, and 3 E**). The state of São Paulo was pointed as the most probable (PSP = 0.68) epicenter of lineage P.1.7 dissemination to several states from Southern (Paraná, Rio Grande do Sul and Santa Catarina), Southeastern (Rio de Janeiro), Central-Western (Goiás and Mato Grosso do Sul), Northeastern (Alagoas, Maranhão, Paraíba, and Pernambuco), and Northern (Pará and Tocantins) Brazilian regions (**Figure 3 D**). We found evidence of large local transmission clusters of lineage P.1.7 in Goiás and Maranhão and of secondary disseminations from Goiás to Maranhão, Tocantins, Pará, Paraná and Santa Catarina, and from Maranhão to Amazonas, Pará, Paraná, Goiás and Rio de Janeiro. The state of Rio de Janeiro was pointed as the most probable epicenter (*PSP* = 1) of lineage P.1.8. Despite this variant’s overall low estimated prevalence, we found evidence of its dissemination to the Amazonas and Santa Catarina states and of subsequent local transmission in Santa Catarina (**Figure 3 E**).

### Epidemic expansion of lineages P.1.4 and P.1.6

The increasing relative frequency of lineages P.1.4 and P.1.6 suggest that those variants displayed an effective reproduction number (Re) >1 in the Amazonas state between March and July 2021. To confirm this hypothesis, we used the birth-death skyline (BDSKY) model to estimate the Re of lineages P.1, P.1.4, and P.1.6. Although the high posterior density intervals (HPD) were relatively high, the trajectory of the median Re values was consistent with each lineage’s relative prevalence changes (**Figure 4**). Lineage P.1.4 displayed a roughly constant median Re > 1 (1.08-1.12) overall the study period; while lineage P.1.6 displayed a median Re > 1 (1.09-1.25) from February to early June, but decreased to 0.91 in June-July, coinciding with the drop of its relative frequency from 37.1% in June to 13.5% in early July. Lineage P.1 displayed a median Re > 1 (1.96) during November-December 2020, a Re ∼ 1 during January-March 2021, and a Re < 1 (0.77-0.95) from April to July 2021. In June-July 2021, the ratio Re(P.1.4)/Re(P.1) was 1.41-1.42 and the ratio Re(P.1.6)/Re(P.1) was 1.17-1.38.

**Figure 4.**
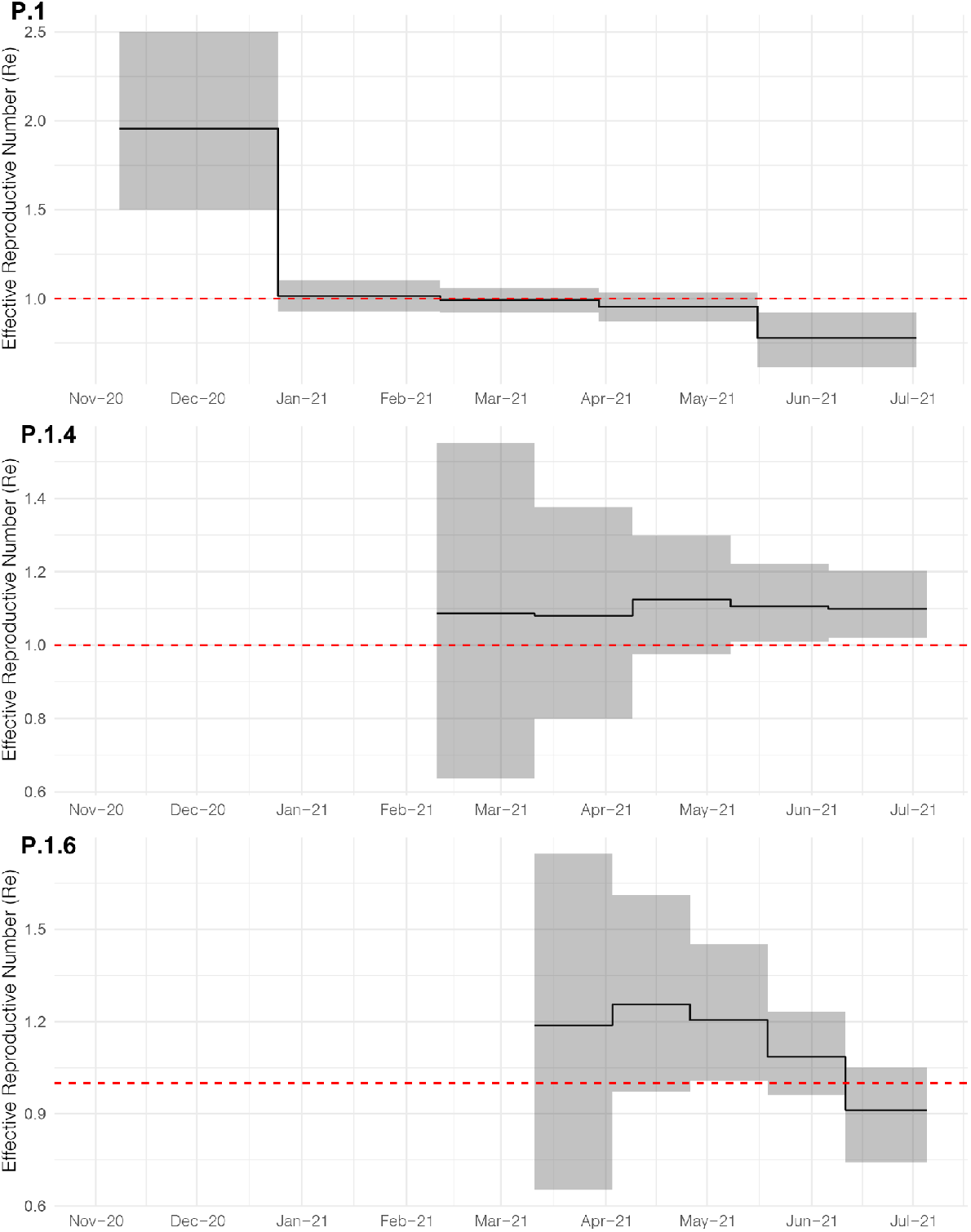
Epidemic trajectories of major SARS-CoV-2 lineages circulating in Amazonas in 2021. Graphs depicting the temporal variation in Re (median and 95%HPD) of Amazonian lineages P.1, P.1.4 and P.1.6 were estimated using the BDSKY approach.

### The spread of P.1+ variants was associated with increasing community viral load

The distribution of real-time RT–PCR cycle threshold (Ct) scores from single or successive cross-sectional samples is a good proxy for virus load in the underlying population and may be shaped by changes in both the infecting virus variant and the epidemic trajectory (8). To test if individuals infected with P.1+ variants might have a higher virus load in the upper respiratory tract than those infected with P.1, we follow-up the evolution of Ct values among SARS-CoV-2 cases diagnosed in Amazonas during the endemic phase of transmission (March to July 2021) when the number of SARS-CoV-2 cases remained roughly stable. Our analyses revealed that as the relative prevalence of P.1+ lineages increase, the mean Ct value of SARS-CoV-2 positive cases progressively reduced from 26.8 (95% CI: 26.5-27.0) in March to 24.4 (95% CI: 24.2-24.7) in July, which correspond to a ∼6-fold increase in the mean viral load of infected subjects (**Figure 5A**). Median Ct values in June-July 2021 were significantly (P <0.0001) lower than in March-May 2021 (**Figure 5 B**).

**Figure 5.**
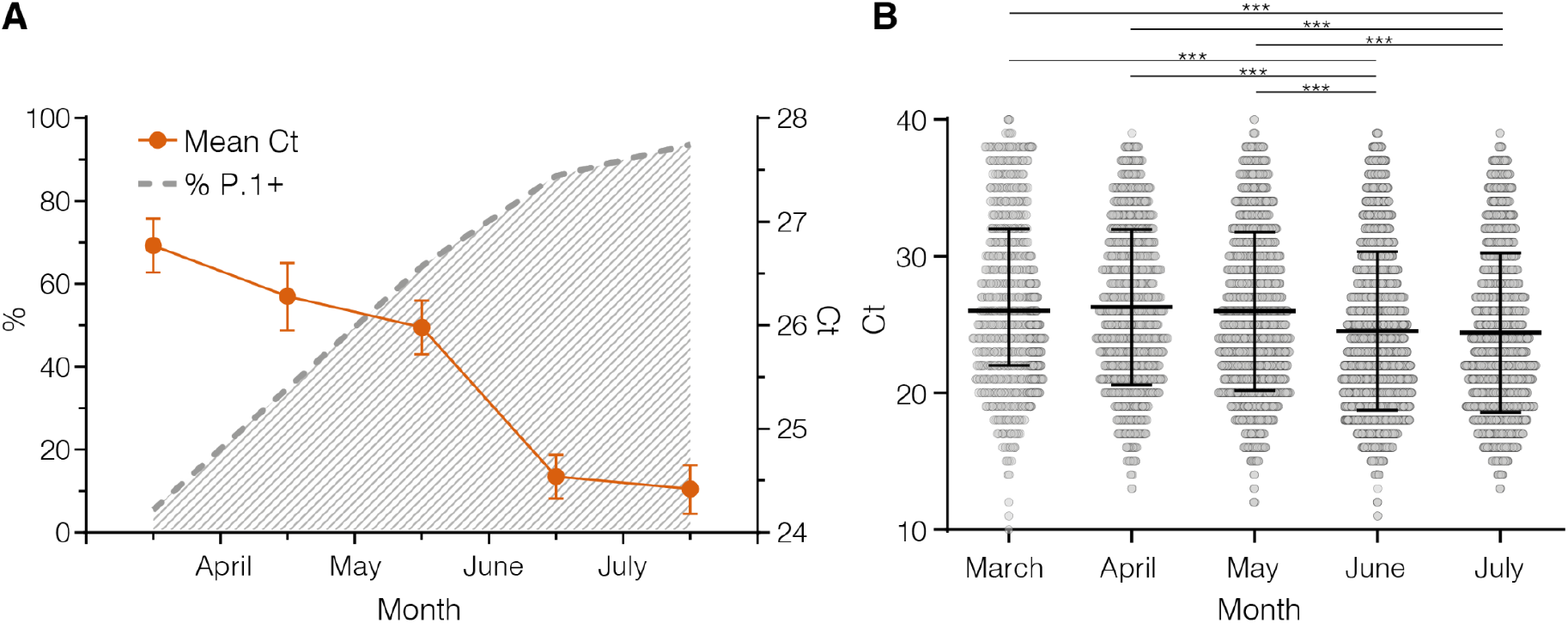
Estimation by RT–PCR of viral load in the upper respiratory tract of SARS-CoV-2 infected patients in Amazonas. **A**, Graph depicting the relative prevalence of P.1+ lineages estimated from whole-genome sequencing (dashed gray line) and the Ct (mean and 95% Confidence Interval) among SARS-CoV-2 positive case**s** (solid line) in Amazonas between March and July 2021. **B**, Comparison of Ct values from March to July 2021. Horizontal bars represent Ct medians and IQR. Two-sided *P-values* for the nonparametric Mann–Whitney test are shown for each group. Two-sided *P-values* <0.05 were considered statistically significant.

### The potential impact of S1/S2 mutations on Spike cleavability

The S1/S2 furin cleavage site (681PRRAR|S686), which is found in a loop region between residues 675-692, is of great importance for SARS-CoV-2 Spike protein structural changes during the initial steps of host cell infection and that influence the invasion success and transmission of the virus (9, 10). The specificity of furin for a polybasic substrate is partly due to the high negative charge distributed close to the active site and mutations S:P681H/R, that exchange neutral/non-polar residues with basic amino acids close to the cleavage site, enhance the S1/S2 cleavability (11-15). We hypothesize that mutation S:N679K may also benefit the enzyme-substrate coupling. To test this hypothesis, metadynamics simulations that modeled the dissociation of the substrate-enzyme complex were used to estimate the relative binding affinity between the wildtype (WT)/mutant peptides mimicking the S protein loop and the furin enzyme. Our analyses predict that replacement of a non-polar residue (P) by a polar one (H) provides a modest increase in affinity as compared to the WT peptide, whereas having a positively charged residue at either position 679 (K) or 681 (R) leads to a dramatic enhancement in binding affinity to the furin binding site (**Figure 6**).

**Figure 6.**
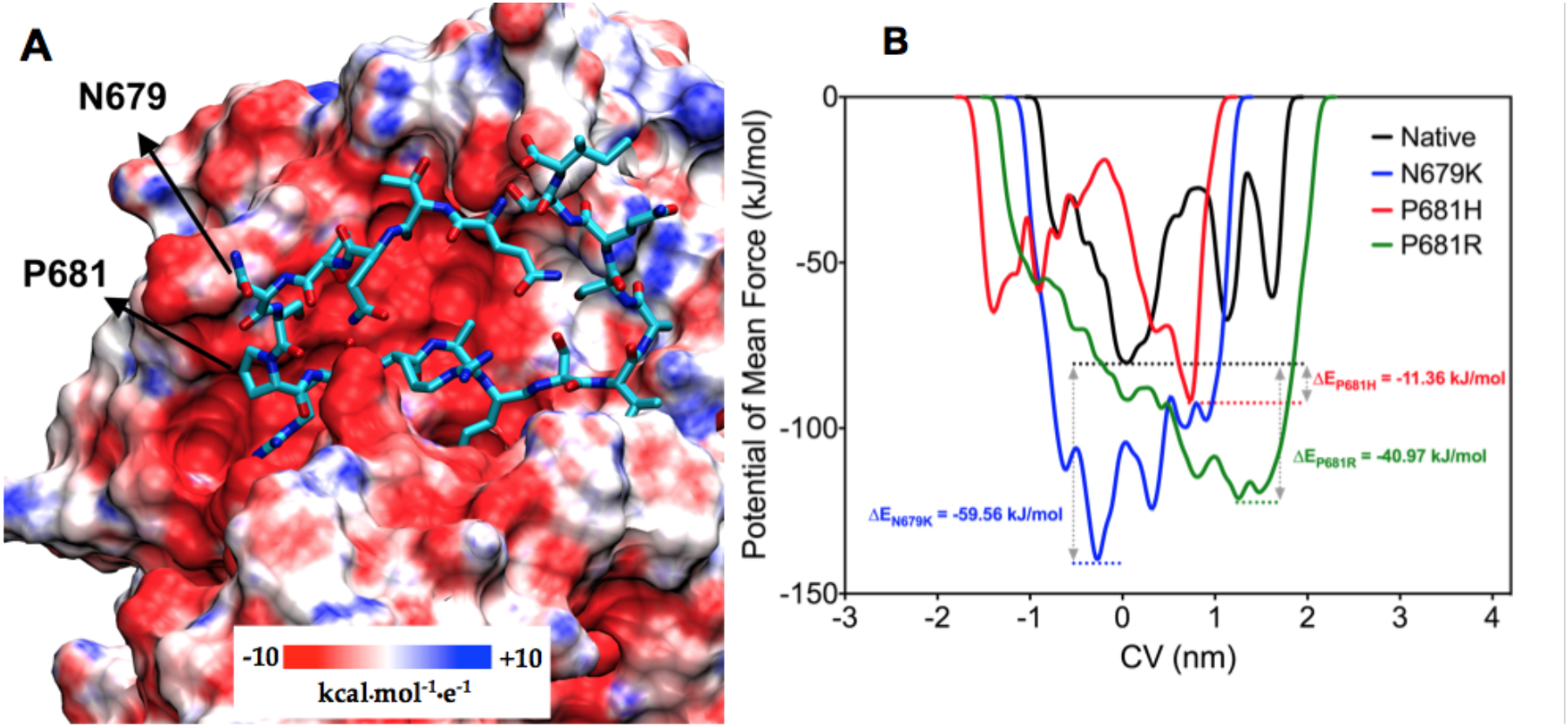
Binding between furin enzyme and the structural motif _679_NSPRRARS_686_ of the SARS-CoV-2 Spike protein. (A) Representation of furin as electrostatic potential surface, showing the negative charge distribution around the loop substrate (represented in licorice cyan). Arrows indicate the native position of variant mutations N679K and P681H/R. (B) Free energy surface landscape of dissociation of modeled loops from furin enzyme depicted through the potential of mean force as a function of the chosen CV. Dashed lines represent the lowest energy basin for the dissociation of the peptides.

### Frequency of P.1+ lineages among vaccine breakthrough cases

The expansion of P.1+ lineages in the Amazonas state may be partly associated with their higher ability to infect individuals who acquired immunity through vaccinations. To test this hypothesis, we compared the frequency of distinct P.1 variants in 38 fully vaccinated breakthrough cases (documented infection occurring 14 days after the second dose) and 104 unvaccinated individuals diagnosed in the Amazonas between April and July 2021. Our analyses revealed that P.1+ lineages were not overrepresented among breakthrough SARS-CoV-2 infections (71%) compared with unvaccinated controls (93%) infected in the same period (**Figure 7**). Significant difference (*P*< 0.0001) in the overall distribution of P.1+ lineages among groups was mainly explained by the much higher frequency of lineage P.1.6 in the unvaccinated (63%) than in the fully vaccinated (32%) group. Although this may indicate a higher susceptibility of unvaccinated individuals to infection with lineage P.1.6 with respect to the fully-vaccinated ones, this is a limited sample size and observations should be interpreted with caution.

**Figure 7.**
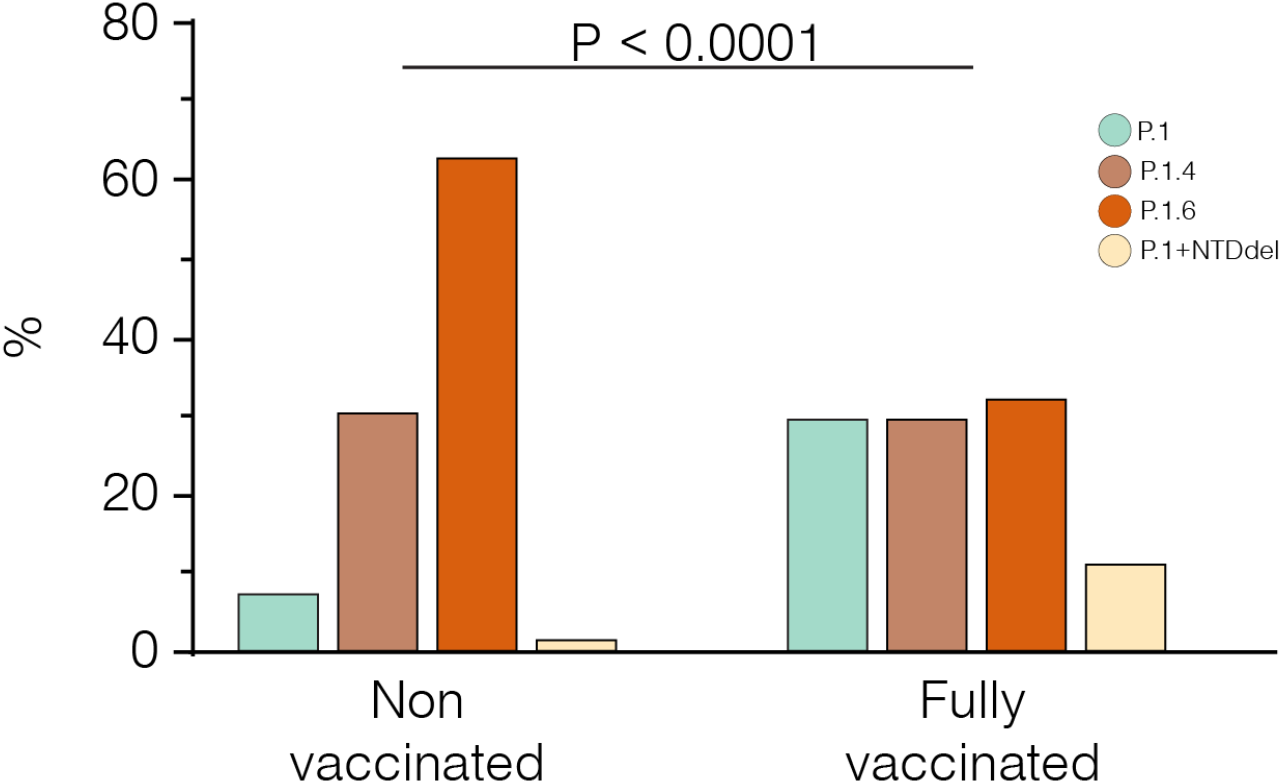
Relative prevalence of P.1 and P.1+ lineages among unvaccinated and fully-vaccinated groups. All breakthrough cases were detected among individuals fully vaccinated with CoronaVac. The *P*-value for the Chi–Square test is shown. *P*-values <0.05 were considered statistically significant.

## Discussion

Understanding how SARS-CoV-2 persists in settings where a high proportion of the population have developed immunity against SARS-CoV-2, due to natural infection or vaccination, has important public health implications in the management of SARS-CoV-2 infections. In this study, we demonstrate that the endemic transmission of SARS-CoV-2 after the second COVID-19 epidemic wave in Amazonas in 2021 was associated with the continuous evolution of the VOC P.1 (Gamma) through the acquisition of NTD deletions (mainly **Δ**144 and **Δ**141-44) or, more frequently, of S1/S2 mutations (P681H and N679K) in the Spike protein. The stable number of SARS-CoV-2 cases registered in the Amazonas between May and July 2021 resulted from two divergent underlying viral dynamics, the rise of the P.1+ variants and the concurrent extinction of the original lineage P.1.

Evidence supports that new P.1+ lineages emerging in Amazonas are more transmissible than the parental P.1 lineage. First, the main P.1+ lineages probably arose in the Amazonas state between mid-February and late April 2021, and their combined prevalence among SARS-CoV-2 positive cases that underwent genomic sequencing increased from 6% in March to >85% in June-July 2021. Second, while lineages P.1.4 and P.1.6 expanded (median Re >1) from March to July 2021, the parental lineage P.1 declined (median Re < 1) in the Amazonian population. Third, the increasing prevalence of P.1+ variants between March and June/July 2021, and particularly of lineages P.1.4 and P.1.6, was correlated with a 6-fold increase in the level of SARS-CoV-2 RNA estimated from the median Ct of positive samples in the Amazonas, suggesting that P.1+-infected adult individuals could be more infectious than those harboring the parental P.1 viruses. It will be essential to determine if these P.1+ lineages are also associated with a distinct clinical evolution.

P.1+ lineages with convergent mutations/deletions also appeared independently in other Brazilian states. A previous study conducted by our group detected the concurrent emergence of multiple P.1+ lineages bearing NTD deletions (including **Δ**144 and **Δ**141-44) in several Brazilian states (7) and the present molecular survey identified the emergence of P.1 lineages with mutations S:N679K and S:P681H/R outside the Amazonas. The major P.1+ lineages circulating outside the Amazonas state were P.1.7 (P.1+P681H), which probably arose in the state of São Paulo around mid-February 2021 and is currently detected in at least ten different states from all country regions; and P.1.8 (P.1+P681R), that probably arose in the state of Rio de Janeiro in early May 2021 and spread to the Southern (Santa Catarina) and Northern (Amazonas) regions. Although it is too early to determine whether these P.1+ lineages will become dominant outside the Amazonas state, their recurrent emergence suggests some transmissibility advantages over the P.1 original variant.

Most genetic changes identified in P.1+ lineages also appear in other VOCs and are predicted or known to affect virus infectivity, immune escape, or both. Different NTD deletions present in VOCs Alpha (**Δ**144), Beta (**Δ**241-243), and Delta (**Δ**157-158) confer resistance to neutralizing antibodies (NAb) directed against the NTD antigenic supersite; while the parental VOC Gamma that lack NTD deletions is more sensitive to those NAb (16-18). The two most common NTD deletions in P.1+ Amazonian lineages (**Δ**144 and **Δ**141-144) confer resistance to anti-NTD NAb and have been shown to emerge in vivo in long-term infections following therapy with convalescent plasma (19, 20) and during acute infections following production of autologous anti-NTD antibodies (21). Interestingly, NTD deletions **Δ**141 and **Δ**142 were among the selected forecasted mutations that may contribute to evolution of VOCs according to a recent study (22). Thus, NTD deletions probably represent a primary adaptive mechanism of further immune evasion of the VOC Gamma evolving in Brazil.

Two VOCs (Alpha and Delta) and several VOIs (AV.1, B.1.1.318, B.1.617.1, B.1.617.3, and P.3) have independently acquired mutations S:P681H/R at the multibasic furin motif. Both mutations have been shown to enhance the S1/S2 cleavability by furin-like proteases and mutation S:P681R, but not S:P681H, also enhances viral replication, viral fusion, and cell-cell viral spread *in vitro* when it occurs in the background of other S mutations (11-15). Mutation S:N679K has not been previously associated with VOCs/VOIs, but the change for an additional basic residue close to the furin cleavage site might enhance the S1/S2 cleavability.

Furin is an endoproteinase whose substrate is a polybasic structural motif of the type R-X-K/R-R↓ (where ↓ represents the peptide bond to be cleaved) (23, 24). In the S protein of SARS-CoV-2, furin recognizes the motif 681-PRRAR↓S-686 which is found in a long loop region between residues 675-692. The specificity of furin for a polybasic substrate is due in part to the high negative charge distributed close to the active site (**Figure 6 A**). Thus, our hypothesis is that the binding between furin and the loops of variants P681H, P681R and N679K has greater stability due to the exchange of neutral/non-polar residues for residues with a basic character. To test our hypothesis, metadynamics simulations were used to estimate the binding strength between the wildtype (WT)/mutant peptides mimicking the S protein loop and the furin enzyme. These simulations modeled the dissociation of the substrate-enzyme complex and computed their relative binding affinity. The dissociation was freely conducted along the CVs-defined pathway, providing the potential of mean force (PMF) barrier between the WT complex and the pre-complex states. For metadynamics simulations in which the employed CV is protein – peptide distance, the PMF is equivalent to the free energy of binding (**Δ***G*).

**Figure 6 B** shows the free energy landscape (FEL) for the unbinding mechanism of the WT and mutant peptides. The FEL was described through the PMF, depicting the phase space sampled from the CVs. As it can be seen, the three PMFs display a well-defined energy minimum, and metastable states during the dissociation process. It is worth noting that these values may differ quantitatively if the full S protein head was used. Additionally, the calculated values refer to energies obtained within the determined Gaussian parameters to the set of compounds under study. Thus, these energies should be used as a measure to compare the binding free energies among them, and not as absolute free energies. From the PMF it is inferred that all mutants present more favorable binding energy, and therefore, higher affinity, as compared to the WT peptide. **Figure 6 B** shows that replacement of a non-polar residue by a polar one (P→H) provides a modest increase in affinity, whereas having a positively charged residue at either position, 679 or 681, leads to a dramatic enhancement in binding affinity to the furin binding site. A recent study showed that mutation S:H655Y, characteristic of the VOC P.1, also enhanced S cleavage (25) and might thus function synergistically with mutations S:N679K and S:P681H/R.

Two critical findings support that endemic transmission of lineage P.1 in the Amazonian population selected more infectious variants rather than variants that are more able to evade prior immunity triggered by natural infections or vaccines. First, we found no evidence that new P.1+ lineages circulating in the Amazonas were overrepresented among post-vaccination breakthrough SARS-CoV-2 infections compared with infections among unvaccinated control. Second, the relative prevalence of P.1+ lineages harboring immune-escape NTD deletions increased to 12.4% in May but decreased in June and July 2021, indicating that those variants displayed a transient transmission advantage despite the continuously growing number of immunized individuals in the Amazonas. This pattern is consistent with the hypothesis that the proportion of immune individuals in the Amazonas probably already exceeded the herd immunity threshold of most P.1 variants, except for those more transmissible lineages P.1.4 and P.1.6.

In summary, our study confirms that endemic transmission of SARS-CoV-2 after the second COVID-19 epidemic wave in the Amazonas state has been associated with the continuous evolution of the VOC P.1 through the acquisition of either Spike mutations at the S1/S2 junction (N679K or P681H) or NTD deletions that probably increased viral infectivity or resistance against antiviral immunity. The steady-state level of SARS-CoV-2 cases observed in Amazonas between May and July 2021 resulted from the concurrent expansion of lineages P.1+ and extinction of parental lineage P.1. The spread of lineages P.1+ in the Amazonas state was probably limited by the increasing number of immunized (naturally infected and vaccinated) individuals. These findings highlight the importance of closely monitoring the evolution of SARS-CoV-2 VOCs as they continue to spread in human populations with decreasing density of susceptible hosts and the urgent need to accelerate the vaccination roll-out to reduce further viral transmission and evolution.

## Material and Methods

### SARS-CoV-2 samples and ethical aspects

We analyzed nasopharyngeal and pharyngeal swabs (NPS) collected between 01st January and 06th July 2021 from residents in the Amazonas state positively tested by real-time RT-PCR as a routine diagnostic for SARS-CoV-2. In total, 1,188 samples were submitted to nucleotide sequencing at the Fiocruz/ILMD under the auspices of the Fiocruz COVID-19 Genomic Surveillance Network, the Amazonas State Health Foundation - Dr Rosemary Costa Pinto (FVS-RCP), and the Brazilian Ministry of Health. This study was approved by the Ethics Committee of the Amazonas State University, which waived signed informed consent (CAAE:25430719.6.0000.5016).

### SARS-CoV-2 amplification and sequencing

The SARS-CoV-2 genomes were recovered using a previously described sequencing protocol (26) or a commercial kit Illumina COVIDSeq Test (Illumina), with the following modifications in the original manufacturer instructions. The reverse transcription step was optimized to use half of the volume described in the Illumina COVIDSEQ Test protocol. We also included a longer incubation time (65°C for 5 minutes) and increasing sequential temperature steps 25°C 15’ / 37°C 15’/ 45°C 15’/ 50°C 15’ / 70°C 15’ and 4°C / ∞. With these modifications we were able to sequence samples with Ct higher than 25, around 28-30. We also included custom primers at a final concentration of 20nM to minimize the dropout observed in the primers target regions when only standard Illumina COVIDSeq Test primer set was used. (COVIDSEQ_3732_FNF 5’ - GTTGTTAATGCAGCCAATGTTTACCTTAAA, COVIDSEQ_4186_FNR CAACTTGCTTTTCACTCTTCATTTCCAAA, COVIDSEQ_20883_FNF 5’ TGCTAATTCCATTGTTTGTAGATTTGACACTA, COVIDSEQ_21285_FNR 5’ - CTGAAGTCTTGTAAAAGTGTTCCAGAGG, COVIDSEQ_24259_FNF 5’ - AACATCACTAGGTTTCAAACTTTACTTGCT, COVIDSEQ_24604_FNR 5’ - ATGCAAATCTGGTGGCGTTAAAAAC). The new generation sequencing libraries were clustered with MiSeq Reagent Kit v3 (600-cycles) on 2 × 150 cycles paired-end runs (Illumina) or with NextSeq 1000 on 2 × 50 cycles. FASTQ reads were generated by the Illumina pipeline at BaseSpace (https://basespace.illumina.com). Consensus sequences were generated using DRAGEN COVID LINEAGE 3.5.1 to 3.5.3, according to the most up-to-date version of this app on each sequencing run. Subsequently, we evaluated the consensus files for quality using Nextclade tool v1.5.2 (https://clades.nextstrain.org), those with more than 1% of ambiguities “Ns” had the FASTQ files imported into Geneious Prime 2021 for trimming and assembling using a customized workflow employing BBDuk and BBMap tools (v38.84) and the NC_045512.2 RefSeq as a template with carefully visually inspection. Using both approaches, we generated consensus sequences with mean depth coverage higher than 800X, excluding duplicate reads. Whole-genome SARS-CoV-2 consensus sequences were initially assigned to viral lineages according to the nomenclature proposed by Rambaut et al. (27), using the Pango Lineage web application (https://pangolin.cog-uk.io) and later confirmed using phylogenetic analyses as explained below.

### Maximum Likelihood Phylogenetic Analysis

SARS-COV-2 P.1 sequences obtained here were aligned with high quality (<1% of N) and complete (>29 kb) lineage P.1 Amazonian sequences that were available in the EpiCoV database in the GISAID (https://www.gisaid.org/) on May 31st, 2021. Sequences were aligned using MAFFT v7.475 (28) and then subjected to maximum-likelihood (ML) phylogenetic analysis using IQ-TREE v2.1.2 (29) under the general time-reversible (GTR) model of nucleotide substitution with a gamma-distributed rate variation among sites, four rate categories (G4), a proportion of invariable sites (I) and empirical base frequencies (F) nucleotide substitution model, as selected by the ModelFinder application (30). The branch support was assessed by the approximate likelihood-ratio test based on the Shimodaira– Hasegawa-like procedure (SH-aLRT) with 1,000 replicates. The temporal signal of the P.1 and P.1+ sequences was assessed from the ML tree by performing a regression analysis of the root-to-tip divergence against sampling time using TempEst (31).

### Bayesian Phylogeography Analysis

Bayesian Phylogeography Analysis. We performed a time-scaled Bayesian phylogeographic analysis for SARS-CoV-2 genomes of lineages P.1.3 to P.1.8 plus P.1+**Δ**144 sampled in Brazil using the Bayesian Markov Chain Monte Carlo (MCMC) approach implemented in BEAST 1.10.4 (32) with BEAGLE library v3 (33) to improve computational time. The Bayesian tree was reconstructed using the GTR+F+I+G4 nucleotide substitution model, the non-parametric Bayesian skyline (BSKL) model as the coalescent tree prior (34), a strict molecular clock model with a uniform substitution rate prior (8 × 10^−4^ substitutions/site/year) and a reversible discrete phylogeographic model (35) with a continuous-time Markov chain (CTMC) rate reference prior (36). MCMC was run sufficiently long to ensure convergence (effective sample size [ESS] > 200) in all parameters estimates as assessed in TRACER v1.7 (37). The maximum clade credibility (MCC) tree was summarized with TreeAnnotator v1.10 and visualized using FigTree v1.4.4 (http://tree.bio.ed.ac.uk/software/figtree/).

### Effective Reproductive Number (Re) Estimation

To estimate the Re trajectories of lineages P.1, P.1.4, and P.1.6 through time in the Amazonas, we used the birth-death skyline (BDSKY) model (38) implemented within BEAST 2 v2.6.5 (39). The sampling rate (d) was set to zero for the period before the oldest sample and estimated from the data afterward. The BDSKY prior settings were as follows: Become Uninfectious Rate (exponential, mean = 36); Reproductive Number (log-normal, mean = 0.8, sd = 0.5); Sampling Proportion (beta, alpha = 1, beta = 100). Origin parameter was conditioned to the root height, and the Re was estimated piecewise over five-time intervals. The molecular clock was as in the time-scaled trees analysis, and the HKY+G4+F substitution model was used. MCMC chains were run until all relevant parameters reached ESS > 200, as explained above.

### Modeling the relative binding strength of N679K, P681H and P681R variants to furin

The furin cleavage site in the Spike protein of SARS-CoV-2 is found in a disordered region, exposed to the solvent. Due to the high flexibility of this loop, the experimentally resolved structures lack atomic coordinates in this region, even when mutations are introduced to express the protein in its pre-fusion conformation. Starting from the crystallographic structure of an inhibitor bound to human furin (PDB ID: 6HLB) (40), the 679-NSPRRARS-686 loop region was built analogously in the enzyme-interacting conformation (**Supplementary Figure 1 A**). To mimic the complete loop linked to the Spike protein, the modeled motif was used as a folding seed (rotamers kept fixed) and the remaining loop residues were modeled using the Rosetta Remodel protocol (41) (**Supplementary Figure 1 B**). The final model comprising the residues 675-QTQTNSPRRARSVASQSI-692 (**Supplementary Figure 1 C**) was minimized using the FastRelax protocol of Rosetta (42) using the REF2015 energy function and maintaining the catalytic residues of furin fixed. Constraints to Ca2+ ion and its coordinating residues were applied with SetupMetalsMover (43). The variants N679K, P681H and P681R loops were built by using Point mutant (“pmut”) scan application of Rosetta onto a modeled native loop (44). The resultant mutant structures were then geometry-optimized following the same protocol with the FastRelax application. The relative binding free energies between furin and the peptides mimicking the Spike furin-loop variants were estimated by metadynamics simulations in an explicitly aqueous environment (45) (as detailed in the **Supplementary Information Material**).

### Statistical analysis

Descriptive statistics, tests for normal distribution (D’Agostino & Pearson and Anderson-Darling), and the non-parametric Mann-Whitney test were used to compare the Ct of SARS-CoV-2 RT-PCR positive samples from the upper respiratory tract of patients over time. Only Ct values from samples analyzed with the same RT-PCR diagnostic assay were compared. The Chi-Square test was used for testing the association between the vaccination status and frequency of P.1 variants. The threshold for statistical significance was set to *P* < 0.05. Graphics and statistical analyses were performed using GraphPad v9.02 (Prism Software, United States).

## Data Availability

All the SARS-CoV-2 genomes generated and analyzed in this study are available at the EpiCov database in GISAID (https://www.gisaid.org/). The list of accession IDs may be found in the attached supplementary information material file.

## Declarations

## Acknowledgments

The authors wish to thank all the health care workers and scientists who have worked hard to deal with this pandemic threat, the GISAID team, and all the EpiCoV database’s submitters. The GISAID acknowledgment table containing sequences used in this study is shown in **Supplementary Table 2**. We also appreciate the support of Genomic Coronavirus Fiocruz Network members and the Respiratory Viruses Genomic Surveillance Network of the General Laboratory Coordination (CGLab) of the Brazilian Ministry of Health (MoH), Brazilian Central Laboratory States (LACENs), and the Amazonas surveillance teams for the partnership in the viral surveillance in Brazil. Funding support FAPEAM (PCTI-EmergeSaude/AM call 005/2020; Rede Genômica de Vigilância em Saúde - REGESAM e COVACManaus); Conselho Nacional de Desenvolvimento Científico e Tecnológico (grant 403276/2020-9); Inova Fiocruz/Fundação Oswaldo Cruz (Grant VPPCB-007-FIO-18-2-30 - Geração de conhecimento); and Departamento de Ciência e Tecnologia (DECIT), Brazilian Ministry of Health. Computer allocation was partly granted by the Brazilian National Scientific Computing Center (LNCC). FGN, GLW, RDL, and GB are supported by the CNPq through their productivity research fellowships (306146/2017-7, 303902/2019-1, 425997/2018-9 and 302317/2017-1 respectively). GB is also funded by the Fundação Carlos Chagas Filho de Amparo à Pesquisa do Estado do Rio de Janeiro – FAPERJ (Grant number E-26/202.896/2018). ED is funded by Fundação de Apoio à Pesquisa do Espírito Santo - FAPES and Conselho Nacional de Desenvolvimento Científico e Tecnológico - CNPq (Decit-SCTIE-MS - SUS Research Project PPSUS - Grant number 157/2021).

## Author contributions

FGN and MMS contributed to laboratory management and obtaining financial support. VN, VS, ALC, FN, GS, MJ, AC, DD, KP, MJB, LG, ELRB, ALAM, ND, LCGP, FV, FM, FBSD, ERS, and PCR contributed to diagnostics and sequencing analysis. CF, TM, LA, JHS, AM, FMC, FFV, GCM, MXS, VSS, MPM and MVL contributed to patient and public health surveillance data. DC, MF and RL contributed to atomistic simulations. GLW, ED, TG, MMS, and PCR participated in study design and contributed to formal data analysis. FGN and GB conceived and designed the study and contributed to data analysis. FGN and GB wrote the first draft and all authors contributed and approved the final manuscript.

## Data availability

All the SARS-CoV-2 genomes generated and analyzed in this study are available at the EpiCov database in GISAID (https://www.gisaid.org/). The list of accession IDs may be found in the attached **supplementary information material** file.

