## Supplementary Table 1 for "Spread of Gamma (P.1) sub-lineages carrying Spike mutations close to the furin cleavage site and deletions in the N-terminal domain drives ongoing transmission of SARS-CoV-2 in Amazonas, Brazil"

**Supplementary Table 1.** Spike Mutations in 1,188 genomes of Gamma or Gamma-like lineages from Amazonas, Brazil (1st January to 6th July 2021).

| | JANUARY N = 96 | | | | FEBRUARY N = 104 | | | | MARCH N = 164 | | | | APRIL N = 167 | | | | MAY N = 286 | | | | JUNE N = 319 | | | | JULY N = 52 | | Total N = 1188 | Total |
|---|---|---|---|---|---|---|---|---|---|---|---|---|---|---|---|---|---|---|---|---|---|---|---|---|---|---|---|---|
| | 1-15 | | 16-31 | | 1-15 | | 16-28 | | 1-15 | | 16-31 | | 1-15 | | 16-30 | | 1-15 | | 16-31 | | 1-15 | | 16-30 | | 1-6 | | | |
| **AA** | **n** | **%** | **n** | **%** | **n** | **%** | **n** | **%** | **n** | **%** | **n** | **%** | **n** | **%** | **n** | **%** | **n** | **%** | **n** | **%** | **n** | **%** | **n** | **%** | **n** | **%** | **n** | **%** |
| **V3G** | 0 | 0.0 | 0 | 0.0 | 0 | 0.0 | 0 | 0.0 | 0 | 0.0 | 0 | 0.0 | 0 | 0.0 | 1 | 0.9 | 0 | 0.0 | 0 | 0.0 | 0 | 0.0 | 0 | 0.0 | 0 | 0.0 | 1 | 0.1 |
| **L5F** | 1 | 1.8 | 1 | 2.4 | 0 | 0.0 | 1 | 2.0 | 0 | 0.0 | 1 | 1.1 | 0 | 0.0 | 2 | 1.9 | 0 | 0.0 | 2 | 1.2 | 2 | 1.5 | 0 | 0.0 | 1 | 1.9 | 11 | 0.9 |
| **S12F** | 0 | 0.0 | 0 | 0.0 | 0 | 0.0 | 0 | 0.0 | 1 | 1.3 | 0 | 0.0 | 0 | 0.0 | 0 | 0.0 | 0 | 0.0 | 0 | 0.0 | 0 | 0.0 | 2 | 1.1 | 0 | 0.0 | 3 | 0.3 |
| **Q14H** | 0 | 0.0 | 0 | 0.0 | 0 | 0.0 | 0 | 0.0 | 1 | 1.3 | 1 | 1.1 | 0 | 0.0 | 0 | 0.0 | 0 | 0.0 | 1 | 0.6 | 0 | 0.0 | 0 | 0.0 | 0 | 0.0 | 3 | 0.3 |
| **L18F** | 55 | 100.0 | 41 | 100.0 | 54 | 100.0 | 50 | 100.0 | 75 | 100.0 | 89 | 100.0 | 59 | 100.0 | 108 | 100.0 | 122 | 100.0 | 164 | 100.0 | 132 | 100.0 | 187 | 100.0 | 52 | 100.0 | 1188 | 100.0 |
| **T19I** | 0 | 0.0 | 0 | 0.0 | 0 | 0.0 | 0 | 0.0 | 0 | 0.0 | 1 | 1.1 | 0 | 0.0 | 0 | 0.0 | 0 | 0.0 | 0 | 0.0 | 0 | 0.0 | 0 | 0.0 | 0 | 0.0 | 1 | 0.1 |
| **T20N** | 55 | 100.0 | 40 | 97.6 | 54 | 100.0 | 50 | 100.0 | 75 | 100.0 | 87 | 97.8 | 59 | 100.0 | 108 | 100.0 | 122 | 100.0 | 164 | 100.0 | 132 | 100.0 | 187 | 100.0 | 52 | 100.0 | 1185 | 99.7 |
| **T22I** | 0 | 0.0 | 0 | 0.0 | 0 | 0.0 | 0 | 0.0 | 0 | 0.0 | 1 | 1.1 | 0 | 0.0 | 0 | 0.0 | 0 | 0.0 | 0 | 0.0 | 0 | 0.0 | 0 | 0.0 | 0 | 0.0 | 1 | 0.1 |
| **P25L** | 0 | 0.0 | 0 | 0.0 | 0 | 0.0 | 0 | 0.0 | 0 | 0.0 | 0 | 0.0 | 0 | 0.0 | 0 | 0.0 | 1 | 0.8 | 0 | 0.0 | 0 | 0.0 | 0 | 0.0 | 0 | 0.0 | 1 | 0.1 |
| **P25S** | 0 | 0.0 | 0 | 0.0 | 0 | 0.0 | 0 | 0.0 | 0 | 0.0 | 0 | 0.0 | 1 | 1.7 | 0 | 0.0 | 0 | 0.0 | 0 | 0.0 | 0 | 0.0 | 0 | 0.0 | 0 | 0.0 | 1 | 0.1 |
| **P26S** | 55 | 100.0 | 41 | 100.0 | 54 | 100.0 | 50 | 100.0 | 75 | 100.0 | 89 | 100.0 | 59 | 100.0 | 108 | 100.0 | 122 | 100.0 | 164 | 100.0 | 132 | 100.0 | 187 | 100.0 | 52 | 100.0 | 1188 | 100.0 |
| **T29I** | 0 | 0.0 | 0 | 0.0 | 0 | 0.0 | 0 | 0.0 | 0 | 0.0 | 0 | 0.0 | 0 | 0.0 | 0 | 0.0 | 0 | 0.0 | 0 | 0.0 | 0 | 0.0 | 1 | 0.5 | 0 | 0.0 | 1 | 0.1 |
| **V42I** | 0 | 0.0 | 0 | 0.0 | 0 | 0.0 | 0 | 0.0 | 0 | 0.0 | 0 | 0.0 | 0 | 0.0 | 0 | 0.0 | 0 | 0.0 | 1 | 0.6 | 0 | 0.0 | 0 | 0.0 | 0 | 0.0 | 1 | 0.1 |
| **H49Y** | 0 | 0.0 | 0 | 0.0 | 0 | 0.0 | 0 | 0.0 | 1 | 1.3 | 3 | 3.4 | 2 | 3.4 | 8 | 7.4 | 9 | 7.4 | 1 | 0.6 | 0 | 0.0 | 0 | 0.0 | 0 | 0.0 | 24 | 2.0 |
| **H49L** | 0 | 0.0 | 0 | 0.0 | 0 | 0.0 | 0 | 0.0 | 0 | 0.0 | 1 | 1.1 | 0 | 0.0 | 0 | 0.0 | 0 | 0.0 | 0 | 0.0 | 0 | 0.0 | 0 | 0.0 | 0 | 0.0 | 1 | 0.1 |
| **A67S** | 0 | 0.0 | 0 | 0.0 | 0 | 0.0 | 0 | 0.0 | 0 | 0.0 | 0 | 0.0 | 0 | 0.0 | 2 | 1.9 | 2 | 1.6 | 1 | 0.6 | 3 | 2.3 | 3 | 1.6 | 1 | 1.9 | 12 | 1.0 |
| **A67T** | 0 | 0.0 | 0 | 0.0 | 0 | 0.0 | 0 | 0.0 | 0 | 0.0 | 0 | 0.0 | 0 | 0.0 | 1 | 0.9 | 0 | 0.0 | 0 | 0.0 | 0 | 0.0 | 0 | 0.0 | 0 | 0.0 | 1 | 0.1 |
| **I68V** | 0 | 0.0 | 0 | 0.0 | 0 | 0.0 | 0 | 0.0 | 0 | 0.0 | 0 | 0.0 | 0 | 0.0 | 0 | 0.0 | 2 | 1.6 | 0 | 0.0 | 0 | 0.0 | 0 | 0.0 | 0 | 0.0 | 2 | 0.2 |

| | | | | | | | | | | | | | | | | | | | | | | | | | | | | |
|---|---|---|---|---|---|---|---|---|---|---|---|---|---|---|---|---|---|---|---|---|---|---|---|---|---|---|---|---|
| **T95I** | 0 | 0.0 | 0 | 0.0 | 1 | 1.9 | 0 | 0.0 | 0 | 0.0 | 0 | 0.0 | 0 | 0.0 | 0 | 0.0 | 0 | 0.0 | 0 | 0.0 | 0 | 0.0 | 0 | 0.0 | 0 | 0.0 | 1 | 0.1 |
| **S98F** | 0 | 0.0 | 0 | 0.0 | 0 | 0.0 | 0 | 0.0 | 0 | 0.0 | 0 | 0.0 | 0 | 0.0 | 0 | 0.0 | 1 | 0.8 | 0 | 0.0 | 0 | 0.0 | 0 | 0.0 | 0 | 0.0 | 1 | 0.1 |
| **D138Y** | 55 | 100.0 | 41 | 100.0 | 51 | 94.4 | 49 | 98.0 | 75 | 100.0 | 89 | 100.0 | 59 | 100.0 | 107 | 99.1 | 121 | 99.2 | 163 | 99.4 | 132 | 100.0 | 187 | 100.0 | 52 | 100.0 | 1181 | 99.4 |
| **L141F** | 0 | 0.0 | 0 | 0.0 | 0 | 0.0 | 0 | 0.0 | 0 | 0.0 | 0 | 0.0 | 0 | 0.0 | 0 | 0.0 | 2 | 1.6 | 0 | 0.0 | 0 | 0.0 | 0 | 0.0 | 0 | 0.0 | 2 | 0.2 |
| **Y144D** | 0 | 0.0 | 0 | 0.0 | 1 | 1.9 | 0 | 0.0 | 0 | 0.0 | 0 | 0.0 | 0 | 0.0 | 0 | 0.0 | 0 | 0.0 | 0 | 0.0 | 0 | 0.0 | 0 | 0.0 | 0 | 0.0 | 1 | 0.1 |
| **H146Y** | 0 | 0.0 | 0 | 0.0 | 0 | 0.0 | 0 | 0.0 | 0 | 0.0 | 0 | 0.0 | 0 | 0.0 | 0 | 0.0 | 0 | 0.0 | 0 | 0.0 | 0 | 0.0 | 1 | 0.5 | 0 | 0.0 | 1 | 0.1 |
| **W152L** | 0 | 0.0 | 0 | 0.0 | 5 | 9.3 | 0 | 0.0 | 0 | 0.0 | 0 | 0.0 | 0 | 0.0 | 0 | 0.0 | 0 | 0.0 | 0 | 0.0 | 0 | 0.0 | 0 | 0.0 | 0 | 0.0 | 5 | 0.4 |
| **W152C** | 0 | 0.0 | 0 | 0.0 | 0 | 0.0 | 0 | 0.0 | 0 | 0.0 | 0 | 0.0 | 0 | 0.0 | 0 | 0.0 | 1 | 0.8 | 0 | 0.0 | 0 | 0.0 | 0 | 0.0 | 0 | 0.0 | 1 | 0.1 |
| **M153I** | 0 | 0.0 | 0 | 0.0 | 0 | 0.0 | 0 | 0.0 | 0 | 0.0 | 0 | 0.0 | 0 | 0.0 | 1 | 0.9 | 0 | 0.0 | 0 | 0.0 | 0 | 0.0 | 0 | 0.0 | 0 | 0.0 | 1 | 0.1 |
| **N165D** | 0 | 0.0 | 0 | 0.0 | 0 | 0.0 | 0 | 0.0 | 0 | 0.0 | 0 | 0.0 | 0 | 0.0 | 0 | 0.0 | 0 | 0.0 | 1 | 0.6 | 0 | 0.0 | 0 | 0.0 | 0 | 0.0 | 1 | 0.1 |
| **P174S** | 0 | 0.0 | 0 | 0.0 | 0 | 0.0 | 0 | 0.0 | 0 | 0.0 | 0 | 0.0 | 0 | 0.0 | 0 | 0.0 | 0 | 0.0 | 0 | 0.0 | 0 | 0.0 | 1 | 0.5 | 0 | 0.0 | 1 | 0.1 |
| **L176F** | 0 | 0.0 | 0 | 0.0 | 0 | 0.0 | 0 | 0.0 | 0 | 0.0 | 0 | 0.0 | 1 | 1.7 | 0 | 0.0 | 0 | 0.0 | 0 | 0.0 | 0 | 0.0 | 0 | 0.0 | 0 | 0.0 | 1 | 0.1 |
| **K182R** | 0 | 0.0 | 0 | 0.0 | 0 | 0.0 | 0 | 0.0 | 0 | 0.0 | 1 | 1.1 | 0 | 0.0 | 0 | 0.0 | 0 | 0.0 | 0 | 0.0 | 0 | 0.0 | 0 | 0.0 | 0 | 0.0 | 1 | 0.1 |
| **R190S** | 53 | 96.4 | 40 | 97.6 | 54 | 100.0 | 50 | 100.0 | 75 | 100.0 | 89 | 100.0 | 59 | 100.0 | 108 | 100.0 | 122 | 100.0 | 163 | 99.4 | 132 | 100.0 | 187 | 100.0 | 52 | 100.0 | 1184 | 99.7 |
| **V193L** | 0 | 0.0 | 0 | 0.0 | 0 | 0.0 | 0 | 0.0 | 0 | 0.0 | 0 | 0.0 | 0 | 0.0 | 0 | 0.0 | 1 | 0.8 | 0 | 0.0 | 0 | 0.0 | 0 | 0.0 | 0 | 0.0 | 1 | 0.1 |
| **N211S** | 0 | 0.0 | 0 | 0.0 | 0 | 0.0 | 0 | 0.0 | 0 | 0.0 | 0 | 0.0 | 1 | 1.7 | 1 | 0.9 | 1 | 0.8 | 0 | 0.0 | 0 | 0.0 | 0 | 0.0 | 0 | 0.0 | 3 | 0.3 |
| **D215G** | 0 | 0.0 | 0 | 0.0 | 0 | 0.0 | 0 | 0.0 | 1 | 1.3 | 1 | 1.1 | 0 | 0.0 | 4 | 3.7 | 2 | 1.6 | 1 | 0.6 | 0 | 0.0 | 0 | 0.0 | 0 | 0.0 | 9 | 0.8 |
| **D215H** | 0 | 0.0 | 0 | 0.0 | 0 | 0.0 | 0 | 0.0 | 0 | 0.0 | 0 | 0.0 | 0 | 0.0 | 0 | 0.0 | 0 | 0.0 | 0 | 0.0 | 0 | 0.0 | 1 | 0.5 | 0 | 0.0 | 1 | 0.1 |
| **L216F** | 0 | 0.0 | 0 | 0.0 | 0 | 0.0 | 1 | 2.0 | 1 | 1.3 | 0 | 0.0 | 0 | 0.0 | 0 | 0.0 | 0 | 0.0 | 0 | 0.0 | 0 | 0.0 | 0 | 0.0 | 0 | 0.0 | 2 | 0.2 |
| **G232V** | 0 | 0.0 | 0 | 0.0 | 0 | 0.0 | 0 | 0.0 | 1 | 1.3 | 0 | 0.0 | 0 | 0.0 | 0 | 0.0 | 0 | 0.0 | 0 | 0.0 | 0 | 0.0 | 0 | 0.0 | 0 | 0.0 | 1 | 0.1 |
| **N234K** | 0 | 0.0 | 0 | 0.0 | 0 | 0.0 | 0 | 0.0 | 0 | 0.0 | 0 | 0.0 | 0 | 0.0 | 0 | 0.0 | 0 | 0.0 | 1 | 0.6 | 0 | 0.0 | 0 | 0.0 | 0 | 0.0 | 1 | 0.1 |
| **A243S** | 0 | 0.0 | 0 | 0.0 | 0 | 0.0 | 0 | 0.0 | 0 | 0.0 | 1 | 1.1 | 3 | 5.1 | 0 | 0.0 | 0 | 0.0 | 0 | 0.0 | 0 | 0.0 | 0 | 0.0 | 0 | 0.0 | 4 | 0.3 |
| **H245Y** | 0 | 0.0 | 0 | 0.0 | 0 | 0.0 | 0 | 0.0 | 0 | 0.0 | 0 | 0.0 | 0 | 0.0 | 0 | 0.0 | 0 | 0.0 | 0 | 0.0 | 1 | 0.8 | 0 | 0.0 | 0 | 0.0 | 1 | 0.1 |
| **P251L** | 0 | 0.0 | 0 | 0.0 | 0 | 0.0 | 0 | 0.0 | 0 | 0.0 | 0 | 0.0 | 0 | 0.0 | 0 | 0.0 | 0 | 0.0 | 0 | 0.0 | 0 | 0.0 | 0 | 0.0 | 1 | 1.9 | 1 | 0.1 |

| | | | | | | | | | | | | | | | | | | | | | | | | | | | | |
|---|---|---|---|---|---|---|---|---|---|---|---|---|---|---|---|---|---|---|---|---|---|---|---|---|---|---|---|---|
| **S256L** | 0 | 0.0 | 0 | 0.0 | 0 | 0.0 | 0 | 0.0 | 1 | 1.3 | 0 | 0.0 | 0 | 0.0 | 0 | 0.0 | 0 | 0.0 | 0 | 0.0 | 0 | 0.0 | 0 | 0.0 | 0 | 0.0 | 1 | 0.1 |
| **G257D** | 0 | 0.0 | 0 | 0.0 | 0 | 0.0 | 0 | 0.0 | 0 | 0.0 | 0 | 0.0 | 1 | 1.7 | 0 | 0.0 | 0 | 0.0 | 0 | 0.0 | 0 | 0.0 | 1 | 0.5 | 0 | 0.0 | 2 | 0.2 |
| **T259I** | 0 | 0.0 | 0 | 0.0 | 0 | 0.0 | 0 | 0.0 | 1 | 1.3 | 0 | 0.0 | 0 | 0.0 | 0 | 0.0 | 0 | 0.0 | 0 | 0.0 | 0 | 0.0 | 0 | 0.0 | 0 | 0.0 | 1 | 0.1 |
| **T259A** | 0 | 0.0 | 0 | 0.0 | 0 | 0.0 | 0 | 0.0 | 0 | 0.0 | 0 | 0.0 | 0 | 0.0 | 0 | 0.0 | 2 | 1.6 | 0 | 0.0 | 2 | 1.5 | 0 | 0.0 | 0 | 0.0 | 4 | 0.3 |
| **L303F** | 0 | 0.0 | 0 | 0.0 | 0 | 0.0 | 0 | 0.0 | 0 | 0.0 | 0 | 0.0 | 0 | 0.0 | 0 | 0.0 | 0 | 0.0 | 0 | 0.0 | 1 | 0.8 | 0 | 0.0 | 0 | 0.0 | 1 | 0.1 |
| **T323I** | 0 | 0.0 | 0 | 0.0 | 1 | 1.9 | 0 | 0.0 | 0 | 0.0 | 0 | 0.0 | 0 | 0.0 | 0 | 0.0 | 0 | 0.0 | 0 | 0.0 | 0 | 0.0 | 0 | 0.0 | 0 | 0.0 | 1 | 0.1 |
| **A363V** | 0 | 0.0 | 0 | 0.0 | 0 | 0.0 | 0 | 0.0 | 0 | 0.0 | 0 | 0.0 | 0 | 0.0 | 0 | 0.0 | 0 | 0.0 | 0 | 0.0 | 0 | 0.0 | 0 | 0.0 | 1 | 1.9 | 1 | 0.1 |
| **V367F** | 0 | 0.0 | 0 | 0.0 | 0 | 0.0 | 1 | 2.0 | 0 | 0.0 | 0 | 0.0 | 0 | 0.0 | 0 | 0.0 | 0 | 0.0 | 0 | 0.0 | 0 | 0.0 | 0 | 0.0 | 0 | 0.0 | 1 | 0.1 |
| **G413V** | 0 | 0.0 | 0 | 0.0 | 1 | 1.9 | 0 | 0.0 | 0 | 0.0 | 0 | 0.0 | 0 | 0.0 | 0 | 0.0 | 0 | 0.0 | 0 | 0.0 | 0 | 0.0 | 0 | 0.0 | 0 | 0.0 | 1 | 0.1 |
| **K417T** | 55 | 100.0 | 41 | 100.0 | 54 | 100.0 | 50 | 100.0 | 75 | 100.0 | 89 | 100.0 | 59 | 100.0 | 108 | 100.0 | 122 | 100.0 | 164 | 100.0 | 132 | 100.0 | 187 | 100.0 | 52 | 100.0 | 1188 | 100.0 |
| **K444N** | 0 | 0.0 | 0 | 0.0 | 0 | 0.0 | 0 | 0.0 | 1 | 1.3 | 0 | 0.0 | 0 | 0.0 | 0 | 0.0 | 0 | 0.0 | 0 | 0.0 | 0 | 0.0 | 0 | 0.0 | 0 | 0.0 | 1 | 0.1 |
| **T470N** | 0 | 0.0 | 0 | 0.0 | 0 | 0.0 | 0 | 0.0 | 0 | 0.0 | 0 | 0.0 | 0 | 0.0 | 0 | 0.0 | 0 | 0.0 | 0 | 0.0 | 1 | 0.8 | 0 | 0.0 | 0 | 0.0 | 1 | 0.1 |
| **E484K** | 54 | 98.2 | 41 | 100.0 | 54 | 100.0 | 50 | 100.0 | 75 | 100.0 | 89 | 100.0 | 59 | 100.0 | 108 | 100.0 | 122 | 100.0 | 164 | 100.0 | 132 | 100.0 | 187 | 100.0 | 52 | 100.0 | 1187 | 99.9 |
| **S494P** | 0 | 0.0 | 0 | 0.0 | 0 | 0.0 | 0 | 0.0 | 0 | 0.0 | 0 | 0.0 | 0 | 0.0 | 0 | 0.0 | 0 | 0.0 | 0 | 0.0 | 0 | 0.0 | 1 | 0.5 | 0 | 0.0 | 1 | 0.1 |
| **N501Y** | 54 | 98.2 | 41 | 100.0 | 54 | 100.0 | 50 | 100.0 | 75 | 100.0 | 89 | 100.0 | 59 | 100.0 | 108 | 100.0 | 122 | 100.0 | 164 | 100.0 | 132 | 100.0 | 187 | 100.0 | 52 | 100.0 | 1187 | 99.9 |
| **Y508H** | 0 | 0.0 | 0 | 0.0 | 0 | 0.0 | 0 | 0.0 | 0 | 0.0 | 0 | 0.0 | 0 | 0.0 | 1 | 0.9 | 0 | 0.0 | 0 | 0.0 | 0 | 0.0 | 0 | 0.0 | 0 | 0.0 | 1 | 0.1 |
| **K537N** | 0 | 0.0 | 0 | 0.0 | 0 | 0.0 | 0 | 0.0 | 0 | 0.0 | 0 | 0.0 | 0 | 0.0 | 0 | 0.0 | 0 | 0.0 | 2 | 1.2 | 1 | 0.8 | 1 | 0.5 | 0 | 0.0 | 4 | 0.3 |
| **N540S** | 0 | 0.0 | 0 | 0.0 | 0 | 0.0 | 0 | 0.0 | 0 | 0.0 | 1 | 1.1 | 0 | 0.0 | 0 | 0.0 | 0 | 0.0 | 0 | 0.0 | 0 | 0.0 | 0 | 0.0 | 0 | 0.0 | 1 | 0.1 |
| **F543L** | 0 | 0.0 | 0 | 0.0 | 0 | 0.0 | 0 | 0.0 | 0 | 0.0 | 0 | 0.0 | 0 | 0.0 | 0 | 0.0 | 2 | 1.6 | 0 | 0.0 | 0 | 0.0 | 0 | 0.0 | 0 | 0.0 | 2 | 0.2 |
| **T547K** | 0 | 0.0 | 0 | 0.0 | 0 | 0.0 | 0 | 0.0 | 1 | 1.3 | 0 | 0.0 | 0 | 0.0 | 0 | 0.0 | 0 | 0.0 | 0 | 0.0 | 0 | 0.0 | 2 | 1.1 | 0 | 0.0 | 3 | 0.3 |
| **T549I** | 0 | 0.0 | 0 | 0.0 | 0 | 0.0 | 0 | 0.0 | 1 | 1.3 | 0 | 0.0 | 0 | 0.0 | 0 | 0.0 | 0 | 0.0 | 0 | 0.0 | 0 | 0.0 | 0 | 0.0 | 0 | 0.0 | 1 | 0.1 |
| **T553I** | 1 | 1.8 | 0 | 0.0 | 0 | 0.0 | 0 | 0.0 | 0 | 0.0 | 1 | 1.1 | 0 | 0.0 | 0 | 0.0 | 0 | 0.0 | 0 | 0.0 | 0 | 0.0 | 0 | 0.0 | 0 | 0.0 | 2 | 0.2 |
| **E554D** | 0 | 0.0 | 0 | 0.0 | 0 | 0.0 | 0 | 0.0 | 0 | 0.0 | 0 | 0.0 | 0 | 0.0 | 0 | 0.0 | 0 | 0.0 | 0 | 0.0 | 2 | 1.5 | 0 | 0.0 | 0 | 0.0 | 2 | 0.2 |
| **A570V** | 0 | 0.0 | 0 | 0.0 | 0 | 0.0 | 1 | 2.0 | 0 | 0.0 | 0 | 0.0 | 0 | 0.0 | 0 | 0.0 | 0 | 0.0 | 0 | 0.0 | 0 | 0.0 | 0 | 0.0 | 0 | 0.0 | 1 | 0.1 |

| | | | | | | | | | | | | | | | | | | | | | | | | | | | | |
|---|---|---|---|---|---|---|---|---|---|---|---|---|---|---|---|---|---|---|---|---|---|---|---|---|---|---|---|---|
| **T572I** | 0 | 0.0 | 0 | 0.0 | 0 | 0.0 | 0 | 0.0 | 0 | 0.0 | 0 | 0.0 | 0 | 0.0 | 0 | 0.0 | 0 | 0.0 | 0 | 0.0 | 0 | 0.0 | 1 | 0.5 | 0 | 0.0 | 1 | 0.1 |
| **E583G** | 0 | 0.0 | 0 | 0.0 | 0 | 0.0 | 0 | 0.0 | 0 | 0.0 | 0 | 0.0 | 0 | 0.0 | 0 | 0.0 | 0 | 0.0 | 0 | 0.0 | 1 | 0.8 | 0 | 0.0 | 0 | 0.0 | 1 | 0.1 |
| **P589Q** | 0 | 0.0 | 0 | 0.0 | 0 | 0.0 | 0 | 0.0 | 0 | 0.0 | 0 | 0.0 | 0 | 0.0 | 0 | 0.0 | 0 | 0.0 | 0 | 0.0 | 0 | 0.0 | 1 | 0.5 | 0 | 0.0 | 1 | 0.1 |
| **Q607H** | 0 | 0.0 | 0 | 0.0 | 0 | 0.0 | 0 | 0.0 | 0 | 0.0 | 0 | 0.0 | 0 | 0.0 | 0 | 0.0 | 1 | 0.8 | 0 | 0.0 | 0 | 0.0 | 0 | 0.0 | 0 | 0.0 | 1 | 0.1 |
| **Q613H** | 0 | 0.0 | 0 | 0.0 | 0 | 0.0 | 0 | 0.0 | 0 | 0.0 | 0 | 0.0 | 0 | 0.0 | 1 | 0.9 | 0 | 0.0 | 0 | 0.0 | 0 | 0.0 | 0 | 0.0 | 0 | 0.0 | 1 | 0.1 |
| **Q613E** | 0 | 0.0 | 0 | 0.0 | 0 | 0.0 | 0 | 0.0 | 0 | 0.0 | 0 | 0.0 | 0 | 0.0 | 0 | 0.0 | 0 | 0.0 | 0 | 0.0 | 0 | 0.0 | 0 | 0.0 | 2 | 3.8 | 2 | 0.2 |
| **D614G** | 55 | 100.0 | 41 | 100.0 | 54 | 100.0 | 50 | 100.0 | 75 | 100.0 | 89 | 100.0 | 59 | 100.0 | 108 | 100.0 | 122 | 100.0 | 164 | 100.0 | 132 | 100.0 | 187 | 100.0 | 52 | 100.0 | 1188 | 100.0 |
| **E619K** | 0 | 0.0 | 0 | 0.0 | 0 | 0.0 | 0 | 0.0 | 0 | 0.0 | 0 | 0.0 | 1 | 1.7 | 0 | 0.0 | 0 | 0.0 | 0 | 0.0 | 0 | 0.0 | 0 | 0.0 | 0 | 0.0 | 1 | 0.1 |
| **V622F** | 0 | 0.0 | 0 | 0.0 | 0 | 0.0 | 0 | 0.0 | 0 | 0.0 | 0 | 0.0 | 0 | 0.0 | 0 | 0.0 | 0 | 0.0 | 0 | 0.0 | 0 | 0.0 | 1 | 0.5 | 0 | 0.0 | 1 | 0.1 |
| **D627Y** | 0 | 0.0 | 0 | 0.0 | 0 | 0.0 | 0 | 0.0 | 0 | 0.0 | 0 | 0.0 | 0 | 0.0 | 0 | 0.0 | 0 | 0.0 | 0 | 0.0 | 2 | 1.5 | 0 | 0.0 | 0 | 0.0 | 2 | 0.2 |
| **A653V** | 0 | 0.0 | 0 | 0.0 | 0 | 0.0 | 0 | 0.0 | 0 | 0.0 | 1 | 1.1 | 0 | 0.0 | 0 | 0.0 | 0 | 0.0 | 0 | 0.0 | 0 | 0.0 | 1 | 0.5 | 0 | 0.0 | 2 | 0.2 |
| **E654A** | 0 | 0.0 | 0 | 0.0 | 0 | 0.0 | 0 | 0.0 | 0 | 0.0 | 0 | 0.0 | 0 | 0.0 | 0 | 0.0 | 0 | 0.0 | 0 | 0.0 | 0 | 0.0 | 0 | 0.0 | 1 | 1.9 | 1 | 0.1 |
| **H655Y** | 55 | 100.0 | 40 | 97.6 | 54 | 100.0 | 50 | 100.0 | 75 | 100.0 | 89 | 100.0 | 59 | 100.0 | 108 | 100.0 | 122 | 100.0 | 164 | 100.0 | 132 | 100.0 | 187 | 100.0 | 52 | 100.0 | 1187 | 99.9 |
| **S659L** | 0 | 0.0 | 0 | 0.0 | 1 | 1.9 | 0 | 0.0 | 0 | 0.0 | 0 | 0.0 | 0 | 0.0 | 0 | 0.0 | 0 | 0.0 | 0 | 0.0 | 0 | 0.0 | 0 | 0.0 | 0 | 0.0 | 1 | 0.1 |
| **Q675H** | 0 | 0.0 | 0 | 0.0 | 0 | 0.0 | 0 | 0.0 | 0 | 0.0 | 0 | 0.0 | 0 | 0.0 | 2 | 1.9 | 0 | 0.0 | 0 | 0.0 | 2 | 1.5 | 0 | 0.0 | 0 | 0.0 | 4 | 0.3 |
| **N679K** | 0 | 0.0 | 0 | 0.0 | 0 | 0.0 | 0 | 0.0 | 0 | 0.0 | 3 | 3.4 | 7 | 11.9 | 24 | 22.2 | 21 | 17.2 | 53 | 32.3 | 54 | 40.9 | 100 | 53.5 | 40 | 76.9 | 302 | 25.4 |
| **P681H** | 0 | 0.0 | 0 | 0.0 | 0 | 0.0 | 0 | 0.0 | 0 | 0.0 | 1 | 1.1 | 4 | 6.8 | 14 | 13.0 | 25 | 20.5 | 48 | 29.3 | 49 | 37.1 | 61 | 32.6 | 7 | 13.5 | 209 | 17.6 |
| **P681T** | 0 | 0.0 | 0 | 0.0 | 0 | 0.0 | 0 | 0.0 | 0 | 0.0 | 0 | 0.0 | 0 | 0.0 | 1 | 0.9 | 0 | 0.0 | 0 | 0.0 | 0 | 0.0 | 0 | 0.0 | 0 | 0.0 | 1 | 0.1 |
| **P681R** | 0 | 0.0 | 0 | 0.0 | 0 | 0.0 | 0 | 0.0 | 0 | 0.0 | 0 | 0.0 | 0 | 0.0 | 0 | 0.0 | 0 | 0.0 | 0 | 0.0 | 1 | 0.8 | 0 | 0.0 | 0 | 0.0 | 1 | 0.1 |
| **T719I** | 0 | 0.0 | 0 | 0.0 | 0 | 0.0 | 0 | 0.0 | 0 | 0.0 | 0 | 0.0 | 0 | 0.0 | 0 | 0.0 | 0 | 0.0 | 0 | 0.0 | 0 | 0.0 | 1 | 0.5 | 0 | 0.0 | 1 | 0.1 |
| **E725D** | 0 | 0.0 | 0 | 0.0 | 0 | 0.0 | 0 | 0.0 | 0 | 0.0 | 0 | 0.0 | 0 | 0.0 | 2 | 1.9 | 0 | 0.0 | 0 | 0.0 | 0 | 0.0 | 0 | 0.0 | 0 | 0.0 | 2 | 0.2 |
| **T732I** | 0 | 0.0 | 0 | 0.0 | 0 | 0.0 | 0 | 0.0 | 0 | 0.0 | 0 | 0.0 | 0 | 0.0 | 0 | 0.0 | 0 | 0.0 | 0 | 0.0 | 0 | 0.0 | 1 | 0.5 | 0 | 0.0 | 1 | 0.1 |
| **I770V** | 0 | 0.0 | 0 | 0.0 | 0 | 0.0 | 0 | 0.0 | 0 | 0.0 | 0 | 0.0 | 0 | 0.0 | 0 | 0.0 | 0 | 0.0 | 1 | 0.6 | 0 | 0.0 | 0 | 0.0 | 0 | 0.0 | 1 | 0.1 |
| **Q779K** | 0 | 0.0 | 0 | 0.0 | 0 | 0.0 | 0 | 0.0 | 0 | 0.0 | 0 | 0.0 | 0 | 0.0 | 1 | 0.9 | 0 | 0.0 | 0 | 0.0 | 0 | 0.0 | 0 | 0.0 | 0 | 0.0 | 1 | 0.1 |

| | | | | | | | | | | | | | | | | | | | | | | | | | | | | |
|---|---|---|---|---|---|---|---|---|---|---|---|---|---|---|---|---|---|---|---|---|---|---|---|---|---|---|---|---|
| **E780D** | 1 | 1.8 | 0 | 0.0 | 0 | 0.0 | 0 | 0.0 | 0 | 0.0 | 0 | 0.0 | 0 | 0.0 | 0 | 0.0 | 0 | 0.0 | 0 | 0.0 | 0 | 0.0 | 0 | 0.0 | 0 | 0.0 | 1 | 0.1 |
| **P809L** | 0 | 0.0 | 0 | 0.0 | 0 | 0.0 | 0 | 0.0 | 0 | 0.0 | 0 | 0.0 | 0 | 0.0 | 0 | 0.0 | 0 | 0.0 | 0 | 0.0 | 0 | 0.0 | 1 | 0.5 | 0 | 0.0 | 1 | 0.1 |
| **P812L** | 0 | 0.0 | 0 | 0.0 | 1 | 1.9 | 0 | 0.0 | 0 | 0.0 | 0 | 0.0 | 0 | 0.0 | 0 | 0.0 | 0 | 0.0 | 0 | 0.0 | 0 | 0.0 | 0 | 0.0 | 0 | 0.0 | 1 | 0.1 |
| **D839G** | 0 | 0.0 | 0 | 0.0 | 0 | 0.0 | 1 | 2.0 | 0 | 0.0 | 0 | 0.0 | 0 | 0.0 | 0 | 0.0 | 0 | 0.0 | 0 | 0.0 | 0 | 0.0 | 0 | 0.0 | 0 | 0.0 | 1 | 0.1 |
| **A852V** | 0 | 0.0 | 0 | 0.0 | 0 | 0.0 | 0 | 0.0 | 0 | 0.0 | 0 | 0.0 | 0 | 0.0 | 0 | 0.0 | 1 | 0.8 | 1 | 0.6 | 0 | 0.0 | 0 | 0.0 | 0 | 0.0 | 2 | 0.2 |
| **T859I** | 0 | 0.0 | 0 | 0.0 | 0 | 0.0 | 0 | 0.0 | 0 | 0.0 | 1 | 1.1 | 0 | 0.0 | 0 | 0.0 | 0 | 0.0 | 0 | 0.0 | 0 | 0.0 | 0 | 0.0 | 0 | 0.0 | 1 | 0.1 |
| **V860L** | 0 | 0.0 | 0 | 0.0 | 0 | 0.0 | 0 | 0.0 | 0 | 0.0 | 1 | 1.1 | 0 | 0.0 | 0 | 0.0 | 0 | 0.0 | 0 | 0.0 | 0 | 0.0 | 0 | 0.0 | 0 | 0.0 | 1 | 0.1 |
| **I870V** | 0 | 0.0 | 0 | 0.0 | 0 | 0.0 | 0 | 0.0 | 2 | 2.7 | 0 | 0.0 | 0 | 0.0 | 0 | 0.0 | 0 | 0.0 | 0 | 0.0 | 0 | 0.0 | 0 | 0.0 | 0 | 0.0 | 2 | 0.2 |
| **A871V** | 0 | 0.0 | 0 | 0.0 | 0 | 0.0 | 1 | 2.0 | 0 | 0.0 | 0 | 0.0 | 0 | 0.0 | 0 | 0.0 | 0 | 0.0 | 0 | 0.0 | 0 | 0.0 | 0 | 0.0 | 0 | 0.0 | 1 | 0.1 |
| **T883I** | 0 | 0.0 | 0 | 0.0 | 0 | 0.0 | 1 | 2.0 | 0 | 0.0 | 0 | 0.0 | 0 | 0.0 | 0 | 0.0 | 0 | 0.0 | 0 | 0.0 | 0 | 0.0 | 0 | 0.0 | 0 | 0.0 | 1 | 0.1 |
| **S939F** | 0 | 0.0 | 0 | 0.0 | 0 | 0.0 | 0 | 0.0 | 0 | 0.0 | 0 | 0.0 | 0 | 0.0 | 0 | 0.0 | 0 | 0.0 | 0 | 0.0 | 0 | 0.0 | 1 | 0.5 | 0 | 0.0 | 1 | 0.1 |
| **A942V** | 0 | 0.0 | 0 | 0.0 | 0 | 0.0 | 0 | 0.0 | 0 | 0.0 | 0 | 0.0 | 0 | 0.0 | 0 | 0.0 | 0 | 0.0 | 0 | 0.0 | 0 | 0.0 | 1 | 0.5 | 0 | 0.0 | 1 | 0.1 |
| **Q954L** | 0 | 0.0 | 0 | 0.0 | 0 | 0.0 | 0 | 0.0 | 0 | 0.0 | 0 | 0.0 | 0 | 0.0 | 0 | 0.0 | 0 | 0.0 | 0 | 0.0 | 0 | 0.0 | 1 | 0.5 | 1 | 1.9 | 2 | 0.2 |
| **A1020V** | 0 | 0.0 | 0 | 0.0 | 0 | 0.0 | 0 | 0.0 | 0 | 0.0 | 0 | 0.0 | 0 | 0.0 | 0 | 0.0 | 0 | 0.0 | 0 | 0.0 | 1 | 0.8 | 0 | 0.0 | 1 | 1.9 | 2 | 0.2 |
| **T1027I** | 55 | 100.0 | 40 | 97.6 | 54 | 100.0 | 50 | 100.0 | 75 | 100.0 | 89 | 100.0 | 59 | 100.0 | 108 | 100.0 | 122 | 100.0 | 164 | 100.0 | 132 | 100.0 | 187 | 100.0 | 52 | 100.0 | 1187 | 99.9 |
| **A1078S** | 0 | 0.0 | 0 | 0.0 | 0 | 0.0 | 1 | 2.0 | 0 | 0.0 | 0 | 0.0 | 0 | 0.0 | 0 | 0.0 | 0 | 0.0 | 0 | 0.0 | 0 | 0.0 | 0 | 0.0 | 0 | 0.0 | 1 | 0.1 |
| **D1084Y** | 0 | 0.0 | 0 | 0.0 | 0 | 0.0 | 1 | 2.0 | 0 | 0.0 | 0 | 0.0 | 0 | 0.0 | 0 | 0.0 | 0 | 0.0 | 0 | 0.0 | 0 | 0.0 | 0 | 0.0 | 0 | 0.0 | 1 | 0.1 |
| **R1091H** | 0 | 0.0 | 0 | 0.0 | 0 | 0.0 | 0 | 0.0 | 0 | 0.0 | 0 | 0.0 | 0 | 0.0 | 0 | 0.0 | 1 | 0.8 | 1 | 0.6 | 0 | 0.0 | 0 | 0.0 | 0 | 0.0 | 2 | 0.2 |
| **T1117I** | 0 | 0.0 | 0 | 0.0 | 0 | 0.0 | 0 | 0.0 | 1 | 1.3 | 0 | 0.0 | 0 | 0.0 | 0 | 0.0 | 0 | 0.0 | 0 | 0.0 | 0 | 0.0 | 0 | 0.0 | 0 | 0.0 | 1 | 0.1 |
| **D1139H** | 0 | 0.0 | 1 | 2.4 | 0 | 0.0 | 0 | 0.0 | 0 | 0.0 | 0 | 0.0 | 0 | 0.0 | 0 | 0.0 | 0 | 0.0 | 0 | 0.0 | 0 | 0.0 | 0 | 0.0 | 0 | 0.0 | 1 | 0.1 |
| **D1153Y** | 0 | 0.0 | 2 | 4.9 | 0 | 0.0 | 0 | 0.0 | 0 | 0.0 | 0 | 0.0 | 0 | 0.0 | 0 | 0.0 | 0 | 0.0 | 0 | 0.0 | 0 | 0.0 | 0 | 0.0 | 0 | 0.0 | 2 | 0.2 |
| **D1168Y** | 0 | 0.0 | 0 | 0.0 | 0 | 0.0 | 0 | 0.0 | 1 | 1.3 | 0 | 0.0 | 0 | 0.0 | 0 | 0.0 | 0 | 0.0 | 0 | 0.0 | 0 | 0.0 | 0 | 0.0 | 0 | 0.0 | 1 | 0.1 |
| **G1171V** | 0 | 0.0 | 0 | 0.0 | 0 | 0.0 | 0 | 0.0 | 0 | 0.0 | 0 | 0.0 | 0 | 0.0 | 0 | 0.0 | 1 | 0.8 | 2 | 1.2 | 0 | 0.0 | 0 | 0.0 | 0 | 0.0 | 3 | 0.3 |
| **V1176F** | 55 | 100.0 | 41 | 100.0 | 54 | 100.0 | 50 | 100.0 | 75 | 100.0 | 89 | 100.0 | 59 | 100.0 | 108 | 100.0 | 122 | 100.0 | 164 | 100.0 | 132 | 100.0 | 187 | 100.0 | 52 | 100.0 | 1188 | 100.0 |

| | | | | | | | | | | | | | | | | | | | | | | | | | | | | |
|---|---|---|---|---|---|---|---|---|---|---|---|---|---|---|---|---|---|---|---|---|---|---|---|---|---|---|---|---|
| **E1182Q** | 0 | 0.0 | 0 | 0.0 | 0 | 0.0 | 0 | 0.0 | 0 | 0.0 | 0 | 0.0 | 0 | 0.0 | 0 | 0.0 | 0 | 0.0 | 0 | 0.0 | 0 | 0.0 | 0 | 0.0 | 1 | 1.9 | 1 | 0.1 |
| **L1234F** | 0 | 0.0 | 1 | 2.4 | 1 | 1.9 | 0 | 0.0 | 0 | 0.0 | 0 | 0.0 | 0 | 0.0 | 0 | 0.0 | 0 | 0.0 | 0 | 0.0 | 0 | 0.0 | 0 | 0.0 | 0 | 0.0 | 2 | 0.2 |
| **C1235F** | 0 | 0.0 | 0 | 0.0 | 0 | 0.0 | 0 | 0.0 | 0 | 0.0 | 0 | 0.0 | 0 | 0.0 | 0 | 0.0 | 0 | 0.0 | 0 | 0.0 | 1 | 0.8 | 0 | 0.0 | 0 | 0.0 | 1 | 0.1 |
| **S1252F** | 0 | 0.0 | 0 | 0.0 | 0 | 0.0 | 0 | 0.0 | 0 | 0.0 | 1 | 1.1 | 0 | 0.0 | 0 | 0.0 | 0 | 0.0 | 0 | 0.0 | 0 | 0.0 | 0 | 0.0 | 0 | 0.0 | 1 | 0.1 |
| **E1258D** | 0 | 0.0 | 0 | 0.0 | 0 | 0.0 | 0 | 0.0 | 0 | 0.0 | 0 | 0.0 | 2 | 3.4 | 0 | 0.0 | 0 | 0.0 | 0 | 0.0 | 0 | 0.0 | 0 | 0.0 | 0 | 0.0 | 2 | 0.2 |
| **D1259Y** | 0 | 0.0 | 0 | 0.0 | 0 | 0.0 | 0 | 0.0 | 0 | 0.0 | 1 | 1.1 | 0 | 0.0 | 0 | 0.0 | 0 | 0.0 | 0 | 0.0 | 0 | 0.0 | 0 | 0.0 | 0 | 0.0 | 1 | 0.1 |
| **S1261F** | 0 | 0.0 | 0 | 0.0 | 0 | 0.0 | 0 | 0.0 | 0 | 0.0 | 0 | 0.0 | 0 | 0.0 | 0 | 0.0 | 0 | 0.0 | 1 | 0.6 | 0 | 0.0 | 0 | 0.0 | 0 | 0.0 | 1 | 0.1 |

Legend. Spike mutations (AA) are represented by the wild-type amino acid (1-letter code) - residue position - and the mutant amino acid. The total number (N) of genomes generated each month. as well as the number (n) and the percentage (%) of each mutation at different time intervals. are shown. More than one mutant residue was observed in eight positions (P25L or P25S. H49Y or H49L. A67S or A67T. W152L or W152C. D215G or D215H. T259I or T259A. Q613H or Q613E. and P681H or P681T or P681R). Four mutations at the furin cleavage site were observed. N679K in 302 genomes. P681H in 209 genomes. P681R and P681T found in one genome each. The spike mutations N679K and P681H showed a sharp increase from 3.4% (N679K) and 1.1% (P681H) in late March to 76.9% (N679K) and 13.5% (P681H) in early July. Together. genomes carrying furin site mutations (N679K or P681H) represent more than 90% of those sequenced in July.
