## Supplementary Figure 1 for "Spread of Gamma (P.1) sub-lineages carrying Spike mutations close to the furin cleavage site and deletions in the N-terminal domain drives ongoing transmission of SARS-CoV-2 in Amazonas, Brazil"

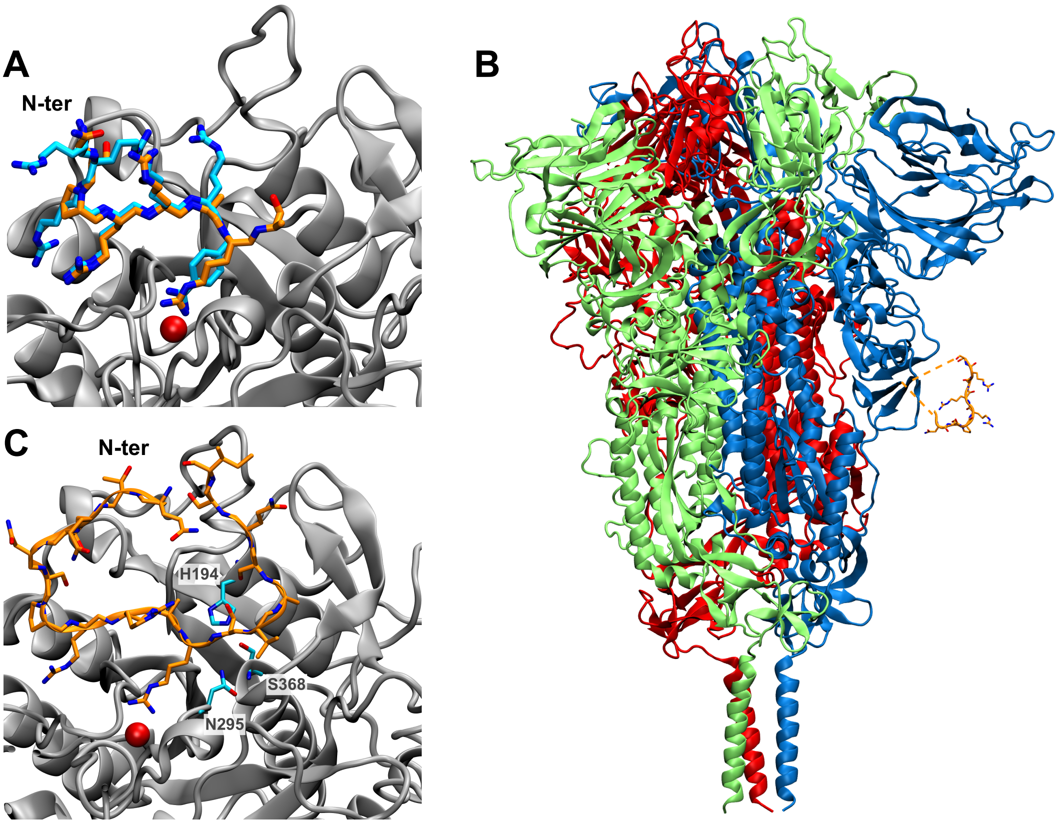


**Figure S1 - (A) Structural motif 679-NSPRRARS-686 (orange, licorice representation) of protein S modeled based on an inhibitor (cyan, licorice representation) that was co-crystallized with the furin enzyme (silver, cartoon representation, $Ca^{2+}$ ion in red) (PDB 6HLB). (B) The modeled 679-NSPRRARS-686 motif (orange, licorice representation) was aligned to the full-Spike homotrimer (chains A, B and C represented in cartoon colors in blue, red and green, respectively) and the remaining residues were modeled to close the loop. (C) Initial input used in calculations with the loop comprising native residues 675-QTQTNSPRRARSVASQSI-692 (orange, licorice representation) in the conformation of interaction with the furin enzyme (cartoon representation, silver). Active site residues His194, Ser368 and Asn295 are shown in licorice representation (nitrogen atoms in blue, carbon atoms in cyan and oxygen atoms in red) and $Ca^{2+}$ ion is represented as a red sphere.**
