## Supplementary Information Material for "Spread of Gamma (P.1) sub-lineages carrying Spike mutations close to the furin cleavage site and deletions in the N-terminal domain drives ongoing transmission of SARS-CoV-2 in Amazonas, Brazil"

#### Atomistic Simulations Additional Information

Prior to the metadynamics simulations, all systems were equilibrated by means of a 5-ns molecular dynamics (MD) simulation. The classical atomistic simulations were performed for the furin enzyme complexed to the wild-type SARS-CoV-2 Spike protein loop, and N679K, P681H and P681R variants, modeled as described above and with the N/C-termini capped. The complexes were embedded in the center of an orthorhombic box with edge dimensions of 2.0 nm of distance from the center of the solute. The systems were solvated with explicit solvent molecules described by the SPC water model[1]. Sodium and chloride ions were added to neutralize each system's total charge while reproducing a buffer of saline solution at 150 nM. The systems were initially energy minimized using 10,000 steps of the steepest descent algorithm. Periodic boundary conditions in were applied in the x, y, and z directions. Holonomic constraints were applied to the bond lengths involving hydrogen atoms in the solute using the LINCS algorithm[2], allowing a 2.0 fs integration time step, in which the leap-frog algorithm was used to integrate the equations of motion. Short-range electrostatics and van der Waals interactions were calculated within the cutoff radius of 1.4 nm. Long-range electrostatics corrections were taken into account by using the reaction-field[3] method beyond a cutoff of 1.4 nm in conjunction with a permittivity dielectric constant of 66. All MD simulations were carried out using the GROMOS parameter set 54A7[4] for the protein, and the GROMOS 53A6[5] parameter set for the ions, within the GROMACS 4.6.7[6]. The equilibration was conducted in the NVT ensemble (constant number of particles, volume, and temperature). The reference temperature was kept at 310 K separately coupling separate v-rescale thermostats[7] for the solute and solvent (electrolytes included) with a relaxation time of 1.0 ps. The systems were previously thermalized by generating initial velocities from a Maxwell-Boltzmann distribution starting at 5 K and progressively increasing to the reference temperature. The simulations were carried out with a 1,000 $kJ.mol^{-1}$ force constant applied to the heavy-backbone atoms of the proteins, allowing only the rearrangement of the water molecules around the solute.

The last frame of the previously equilibrated restrained MD simulations was used as the starting point to the metadynamics. The peptides were steered from their binding sites by defining the collective variables (CV) as the distance between the center of mass of the peptide's α-carbons and the center of mass of the α-carbons of the furin residues interacting initially at 3 Å distant from the peptide. For metadynamics simulations the positional restraints were released, except for the coordinated-$Ca^{2+}$ ions in the furin, in which a 1,000 $kJ.mol^{-1}$ force was applied to restrain their position to the coordinated residues. The exploration of the CV phase space was accomplished by adding Gaussian potentials of height 0.05 kJ/mol and depth of 0.05 nm every 1.0 ps. CVs sampling and calculations were performed for 50 ns via the PLUMED 2.3.5 plugin[8] interfaced with the GROMACS v. 4.6.7. The simulations setup for the metadynamics was the same as described for the restrained MD simulations, save for the inclusion of the Parrinello-Rahman (PR) barostat scheme[9, 10] with a relaxation time of 2.0 ps, leading to simulations in the isothermical-isobaric ensemble (NpT). The choice of PR method for the metadynamics calculations, despite the second-order approach to equilibrium, relies on its correct reproduction of the exact NpT ensemble, which is more appropriate when obtaining thermodynamic quantities. The peptides were detached from the protein along the CV pathway, and the free energy surface (FES) of the process was recursively reconstructed using the sum_hills tool.

### SARS-COV-2 genomes generated in this study

| | | | | |
|---|---|---|---|---|
| EPI_ISL_3050301 | EPI_ISL_2777407 | EPI_ISL_2777474 | EPI_ISL_2777510 | EPI_ISL_2777599 |
| EPI_ISL_2777325 | EPI_ISL_2777408 | EPI_ISL_2777475 | EPI_ISL_2777238 | EPI_ISL_2777600 |
| EPI_ISL_2777615 | EPI_ISL_2777409 | EPI_ISL_2777476 | EPI_ISL_2777511 | EPI_ISL_2777601 |
| EPI_ISL_2777433 | EPI_ISL_1034304 | EPI_ISL_2777477 | EPI_ISL_2777512 | EPI_ISL_2777603 |
| EPI_ISL_2777434 | EPI_ISL_2777410 | EPI_ISL_2777478 | EPI_ISL_2777513 | EPI_ISL_2777604 |
| EPI_ISL_2777436 | EPI_ISL_2777411 | EPI_ISL_2777479 | EPI_ISL_2777357 | EPI_ISL_2777605 |
| EPI_ISL_2777437 | EPI_ISL_2777412 | EPI_ISL_2777480 | EPI_ISL_2777358 | EPI_ISL_2777606 |
| EPI_ISL_2777438 | EPI_ISL_2777413 | EPI_ISL_2777481 | EPI_ISL_2777515 | EPI_ISL_2777609 |
| EPI_ISL_2777439 | EPI_ISL_2777414 | EPI_ISL_2777482 | EPI_ISL_2777516 | EPI_ISL_2777248 |
| EPI_ISL_2777440 | EPI_ISL_2777415 | EPI_ISL_2777483 | EPI_ISL_2777517 | EPI_ISL_2777612 |
| EPI_ISL_2777441 | EPI_ISL_1034306 | EPI_ISL_2777484 | EPI_ISL_2777518 | EPI_ISL_2777616 |
| EPI_ISL_2777443 | EPI_ISL_2777416 | EPI_ISL_2777485 | EPI_ISL_2777989 | EPI_ISL_2777618 |
| EPI_ISL_2777444 | EPI_ISL_2777417 | EPI_ISL_2777486 | EPI_ISL_2777990 | EPI_ISL_2777620 |
| EPI_ISL_2777445 | EPI_ISL_2777418 | EPI_ISL_2777487 | EPI_ISL_2777519 | EPI_ISL_2777621 |
| EPI_ISL_2777446 | EPI_ISL_2777419 | EPI_ISL_2778003 | EPI_ISL_2777520 | EPI_ISL_2777622 |
| EPI_ISL_2777448 | EPI_ISL_2777420 | EPI_ISL_2777488 | EPI_ISL_2777521 | EPI_ISL_2777624 |
| EPI_ISL_2777449 | EPI_ISL_2777421 | EPI_ISL_2777489 | EPI_ISL_2777522 | EPI_ISL_2777627 |
| EPI_ISL_2777450 | EPI_ISL_2777422 | EPI_ISL_2777491 | EPI_ISL_2777992 | EPI_ISL_2777980 |
| EPI_ISL_2777451 | EPI_ISL_2777423 | EPI_ISL_2777491 | EPI_ISL_1068291 | EPI_ISL_2777628 |
| EPI_ISL_2777452 | EPI_ISL_2777424 | EPI_ISL_2778001 | EPI_ISL_2777524 | EPI_ISL_2777630 |
| EPI_ISL_2777490 | EPI_ISL_2777425 | EPI_ISL_2778001 | EPI_ISL_2777525 | EPI_ISL_2777631 |
| EPI_ISL_2777311 | EPI_ISL_2777426 | EPI_ISL_2777492 | EPI_ISL_2777526 | EPI_ISL_2777632 |
| EPI_ISL_2777314 | EPI_ISL_2777427 | EPI_ISL_2777492 | EPI_ISL_2777527 | EPI_ISL_2777634 |
| EPI_ISL_2777547 | EPI_ISL_2777428 | EPI_ISL_2777493 | EPI_ISL_2778002 | EPI_ISL_2777635 |
| EPI_ISL_2777626 | EPI_ISL_2777429 | EPI_ISL_2777494 | EPI_ISL_2777528 | EPI_ISL_2777636 |
| EPI_ISL_2777650 | EPI_ISL_2777430 | EPI_ISL_1068279 | EPI_ISL_2777530 | EPI_ISL_2777637 |
| EPI_ISL_2777678 | EPI_ISL_2777430 | EPI_ISL_1068280 | EPI_ISL_1068292 | EPI_ISL_2777638 |
| EPI_ISL_2777689 | EPI_ISL_2777431 | EPI_ISL_1068281 | EPI_ISL_2777532 | EPI_ISL_2777639 |
| EPI_ISL_2777741 | EPI_ISL_2777431 | EPI_ISL_1068282 | EPI_ISL_2777533 | EPI_ISL_2777641 |
| EPI_ISL_2777851 | EPI_ISL_2777432 | EPI_ISL_1068283 | EPI_ISL_2777534 | EPI_ISL_2777642 |
| EPI_ISL_2777937 | EPI_ISL_2777432 | EPI_ISL_1068284 | EPI_ISL_2777535 | EPI_ISL_2777643 |
| EPI_ISL_2777947 | EPI_ISL_2777995 | EPI_ISL_1068285 | EPI_ISL_2777536 | EPI_ISL_2777645 |
| EPI_ISL_3050601 | EPI_ISL_2777996 | EPI_ISL_2777495 | EPI_ISL_2777537 | EPI_ISL_2777646 |
| EPI_ISL_1533609 | EPI_ISL_2777997 | EPI_ISL_1068286 | EPI_ISL_2777538 | EPI_ISL_2777647 |
| EPI_ISL_2777509 | EPI_ISL_2777998 | EPI_ISL_2778004 | EPI_ISL_2777539 | EPI_ISL_2777651 |
| EPI_ISL_2777356 | EPI_ISL_2777999 | EPI_ISL_2778005 | EPI_ISL_2777540 | EPI_ISL_2777654 |
| EPI_ISL_2777557 | EPI_ISL_2778000 | EPI_ISL_2777239 | EPI_ISL_2777541 | EPI_ISL_2777655 |
| EPI_ISL_2777562 | EPI_ISL_2777435 | EPI_ISL_2777236 | EPI_ISL_2777542 | EPI_ISL_2777657 |
| EPI_ISL_2777564 | EPI_ISL_2777453 | EPI_ISL_2777301 | EPI_ISL_2777543 | EPI_ISL_2777658 |
| EPI_ISL_2777591 | EPI_ISL_1068258 | EPI_ISL_2777240 | EPI_ISL_2777544 | EPI_ISL_2777659 |
| EPI_ISL_2777669 | EPI_ISL_1068260 | EPI_ISL_1068287 | EPI_ISL_2777545 | EPI_ISL_2777662 |
| EPI_ISL_2777690 | EPI_ISL_1068261 | EPI_ISL_1068288 | EPI_ISL_2777546 | EPI_ISL_2777668 |
| EPI_ISL_2777704 | EPI_ISL_1068262 | EPI_ISL_2777241 | EPI_ISL_2777549 | EPI_ISL_2777670 |
| EPI_ISL_2777327 | EPI_ISL_1068263 | EPI_ISL_2777246 | EPI_ISL_2777551 | EPI_ISL_2777675 |
| EPI_ISL_2777765 | EPI_ISL_1068264 | EPI_ISL_2777242 | EPI_ISL_2777553 | EPI_ISL_2777676 |
| EPI_ISL_2777875 | EPI_ISL_1068266 | EPI_ISL_1068289 | EPI_ISL_2777554 | EPI_ISL_2777683 |
| EPI_ISL_2777892 | EPI_ISL_1068267 | EPI_ISL_2777496 | EPI_ISL_2777556 | EPI_ISL_2777686 |
| EPI_ISL_2777898 | EPI_ISL_1068268 | EPI_ISL_2777496 | EPI_ISL_2777558 | EPI_ISL_2777687 |
| EPI_ISL_2777934 | EPI_ISL_1068269 | EPI_ISL_2777496 | EPI_ISL_2777559 | EPI_ISL_2777691 |
| EPI_ISL_2777963 | EPI_ISL_1068270 | EPI_ISL_1068290 | EPI_ISL_2777560 | EPI_ISL_2777692 |
| EPI_ISL_2777299 | EPI_ISL_1068271 | EPI_ISL_2777498 | EPI_ISL_2777561 | EPI_ISL_2777693 |
| EPI_ISL_3050363 | EPI_ISL_1068272 | EPI_ISL_2777499 | EPI_ISL_2777563 | EPI_ISL_2777290 |
| EPI_ISL_3050565 | EPI_ISL_1068273 | EPI_ISL_2777237 | EPI_ISL_2777565 | EPI_ISL_2777250 |
| EPI_ISL_3050558 | EPI_ISL_1068274 | EPI_ISL_2777302 | EPI_ISL_2777285 | EPI_ISL_2777251 |
| EPI_ISL_3050501 | EPI_ISL_1068275 | EPI_ISL_2777304 | EPI_ISL_2777566 | EPI_ISL_2777252 |
| EPI_ISL_3050624 | EPI_ISL_1068276 | EPI_ISL_2777501 | EPI_ISL_2777567 | EPI_ISL_2777253 |
| EPI_ISL_3050387 | EPI_ISL_1068278 | EPI_ISL_2777502 | EPI_ISL_2777569 | EPI_ISL_2777320 |
| EPI_ISL_2777994 | EPI_ISL_2777455 | EPI_ISL_2777503 | EPI_ISL_2777570 | EPI_ISL_2777696 |
| EPI_ISL_2777922 | EPI_ISL_2777456 | EPI_ISL_2777993 | EPI_ISL_2777571 | EPI_ISL_2777254 |
| EPI_ISL_1068268 | EPI_ISL_2777457 | EPI_ISL_2777504 | EPI_ISL_2777572 | EPI_ISL_2777255 |
| EPI_ISL_1068273 | EPI_ISL_2777457 | EPI_ISL_2777505 | EPI_ISL_2777573 | EPI_ISL_2777256 |
| EPI_ISL_2777396 | EPI_ISL_2777458 | EPI_ISL_2102018 | EPI_ISL_2777574 | EPI_ISL_2777697 |
| EPI_ISL_2777397 | EPI_ISL_2777458 | EPI_ISL_2777306 | EPI_ISL_2777575 | EPI_ISL_2777698 |
| EPI_ISL_2777398 | EPI_ISL_2777459 | EPI_ISL_2777506 | EPI_ISL_2777576 | EPI_ISL_2777699 |
| EPI_ISL_2777399 | EPI_ISL_2777459 | EPI_ISL_2777309 | EPI_ISL_2777581 | EPI_ISL_2777257 |
| EPI_ISL_2777401 | EPI_ISL_2777469 | EPI_ISL_2777986 | EPI_ISL_2777584 | EPI_ISL_2777700 |
| EPI_ISL_2777402 | EPI_ISL_2777469 | EPI_ISL_2777987 | EPI_ISL_2777585 | EPI_ISL_2777258 |
| EPI_ISL_2777403 | EPI_ISL_2777470 | EPI_ISL_2777507 | EPI_ISL_2777586 | EPI_ISL_2777259 |
| EPI_ISL_2777404 | EPI_ISL_2777471 | EPI_ISL_2777988 | EPI_ISL_2777588 | EPI_ISL_2777260 |
| EPI_ISL_2777405 | EPI_ISL_2777472 | EPI_ISL_2777313 | EPI_ISL_2777589 | EPI_ISL_2777701 |
| EPI_ISL_2777406 | EPI_ISL_2777473 | EPI_ISL_2777508 | EPI_ISL_2777593 | EPI_ISL_2777292 |

| | | | | |
|---|---|---|---|---|
| EPI_ISL_2777702 | EPI_ISL_2777751 | EPI_ISL_2777847 | EPI_ISL_3050502 | EPI_ISL_4030328 |
| EPI_ISL_2777703 | EPI_ISL_2777752 | EPI_ISL_2777848 | EPI_ISL_3050503 | EPI_ISL_4030310 |
| EPI_ISL_2777261 | EPI_ISL_2777754 | EPI_ISL_2777849 | EPI_ISL_3050337 | EPI_ISL_4030329 |
| EPI_ISL_2777262 | EPI_ISL_2777755 | EPI_ISL_2777850 | EPI_ISL_3050344 | EPI_ISL_4030330 |
| EPI_ISL_2777263 | EPI_ISL_2777757 | EPI_ISL_2777855 | EPI_ISL_3050401 | EPI_ISL_4030331 |
| EPI_ISL_2777264 | EPI_ISL_2777758 | EPI_ISL_2777857 | EPI_ISL_3050519 | EPI_ISL_4030332 |
| EPI_ISL_2777705 | EPI_ISL_2777759 | EPI_ISL_2777858 | EPI_ISL_3050434 | EPI_ISL_4030333 |
| EPI_ISL_2777321 | EPI_ISL_2777762 | EPI_ISL_2777859 | EPI_ISL_3050315 | EPI_ISL_4030334 |
| EPI_ISL_2777706 | EPI_ISL_2777763 | EPI_ISL_2777861 | EPI_ISL_3050354 | EPI_ISL_4030335 |
| EPI_ISL_2777265 | EPI_ISL_2777764 | EPI_ISL_2777862 | EPI_ISL_3050403 | EPI_ISL_4030336 |
| EPI_ISL_2777266 | EPI_ISL_2777766 | EPI_ISL_2777863 | EPI_ISL_3050506 | EPI_ISL_4030337 |
| EPI_ISL_2777267 | EPI_ISL_2777767 | EPI_ISL_2777864 | EPI_ISL_4030320 | EPI_ISL_4030338 |
| EPI_ISL_2777707 | EPI_ISL_2777768 | EPI_ISL_2777867 | EPI_ISL_3050636 | EPI_ISL_4030339 |
| EPI_ISL_2777708 | EPI_ISL_2777340 | EPI_ISL_2777868 | EPI_ISL_4030318 | EPI_ISL_4030340 |
| EPI_ISL_2777268 | EPI_ISL_2777769 | EPI_ISL_2777293 | EPI_ISL_4030305 | EPI_ISL_4030341 |
| EPI_ISL_2777709 | EPI_ISL_2777771 | EPI_ISL_2777870 | EPI_ISL_3050566 | EPI_ISL_4030342 |
| EPI_ISL_2777269 | EPI_ISL_2777772 | EPI_ISL_2777871 | EPI_ISL_3050302 | EPI_ISL_4030343 |
| EPI_ISL_2777270 | EPI_ISL_2777775 | EPI_ISL_2777872 | EPI_ISL_3050303 | EPI_ISL_4030344 |
| EPI_ISL_2777710 | EPI_ISL_2777776 | EPI_ISL_2777873 | EPI_ISL_3050332 | EPI_ISL_4030345 |
| EPI_ISL_2777324 | EPI_ISL_2777779 | EPI_ISL_2777235 | EPI_ISL_3050464 | EPI_ISL_4030346 |
| EPI_ISL_2777711 | EPI_ISL_2777780 | EPI_ISL_2777874 | EPI_ISL_3050309 | EPI_ISL_4030347 |
| EPI_ISL_2777271 | EPI_ISL_2777781 | EPI_ISL_2777876 | EPI_ISL_4030306 | EPI_ISL_4030348 |
| EPI_ISL_2777272 | EPI_ISL_2777782 | EPI_ISL_2777877 | EPI_ISL_3050593 | EPI_ISL_4030349 |
| EPI_ISL_2777273 | EPI_ISL_2777783 | EPI_ISL_2777878 | EPI_ISL_3050319 | EPI_ISL_4030350 |
| EPI_ISL_2777274 | EPI_ISL_2777784 | EPI_ISL_2777879 | EPI_ISL_3050406 | EPI_ISL_4030351 |
| EPI_ISL_2777275 | EPI_ISL_2777785 | EPI_ISL_2777880 | EPI_ISL_3050605 | EPI_ISL_4030352 |
| EPI_ISL_2777328 | EPI_ISL_2777786 | EPI_ISL_2777881 | EPI_ISL_4030321 | EPI_ISL_4030353 |
| EPI_ISL_2777713 | EPI_ISL_2777787 | EPI_ISL_2777882 | EPI_ISL_3050490 | EPI_ISL_4030354 |
| EPI_ISL_2777714 | EPI_ISL_2777788 | EPI_ISL_2777883 | EPI_ISL_4030322 | EPI_ISL_4030315 |
| EPI_ISL_2777715 | EPI_ISL_2777789 | EPI_ISL_2777884 | EPI_ISL_3050407 | EPI_ISL_4030355 |
| EPI_ISL_2777276 | EPI_ISL_2777790 | EPI_ISL_2777885 | EPI_ISL_4030323 | EPI_ISL_4030317 |
| EPI_ISL_2777277 | EPI_ISL_2777791 | EPI_ISL_2777886 | EPI_ISL_3050499 | EPI_ISL_4030308 |
| EPI_ISL_2777278 | EPI_ISL_2777792 | EPI_ISL_2777888 | EPI_ISL_3050312 | EPI_ISL_4030309 |
| EPI_ISL_2777716 | EPI_ISL_2777793 | EPI_ISL_2777889 | EPI_ISL_3050367 | EPI_ISL_4030356 |
| EPI_ISL_2777717 | EPI_ISL_2777794 | EPI_ISL_2777894 | EPI_ISL_3050470 | EPI_ISL_4030316 |
| EPI_ISL_2777279 | EPI_ISL_2777795 | EPI_ISL_2777896 | EPI_ISL_3050471 | EPI_ISL_4030357 |
| EPI_ISL_2777280 | EPI_ISL_2777796 | EPI_ISL_2777897 | EPI_ISL_3050627 | EPI_ISL_4030358 |
| EPI_ISL_2777281 | EPI_ISL_2777797 | EPI_ISL_2777899 | EPI_ISL_3050538 | EPI_ISL_4030359 |
| EPI_ISL_2777718 | EPI_ISL_2102063 | EPI_ISL_2777900 | EPI_ISL_3050570 | EPI_ISL_4030360 |
| EPI_ISL_2777719 | EPI_ISL_2777798 | EPI_ISL_2777284 | EPI_ISL_3050384 | EPI_ISL_4030361 |
| EPI_ISL_2777282 | EPI_ISL_2777247 | EPI_ISL_2777903 | EPI_ISL_3050391 | EPI_ISL_4030362 |
| EPI_ISL_2777283 | EPI_ISL_2777799 | EPI_ISL_2777911 | EPI_ISL_3050394 | EPI_ISL_4030363 |
| EPI_ISL_2777720 | EPI_ISL_2777800 | EPI_ISL_2777913 | EPI_ISL_3050436 | EPI_ISL_4030364 |
| EPI_ISL_2777721 | EPI_ISL_2777801 | EPI_ISL_2777915 | EPI_ISL_4030307 | EPI_ISL_4030365 |
| EPI_ISL_2777722 | EPI_ISL_2777802 | EPI_ISL_2777916 | EPI_ISL_4030312 | EPI_ISL_4030366 |
| EPI_ISL_2777723 | EPI_ISL_2777803 | EPI_ISL_2777917 | EPI_ISL_3050298 | EPI_ISL_4030367 |
| EPI_ISL_2777330 | EPI_ISL_2777804 | EPI_ISL_2777919 | EPI_ISL_3050375 | EPI_ISL_1261123 |
| EPI_ISL_2777331 | EPI_ISL_2777805 | EPI_ISL_2777923 | EPI_ISL_3050376 | EPI_ISL_2777969 |
| EPI_ISL_2777333 | EPI_ISL_2777806 | EPI_ISL_2777932 | EPI_ISL_4030313 | EPI_ISL_2777970 |
| EPI_ISL_2777724 | EPI_ISL_2777807 | EPI_ISL_2777935 | EPI_ISL_4030311 | EPI_ISL_2777971 |
| EPI_ISL_2777725 | EPI_ISL_2777808 | EPI_ISL_2777936 | EPI_ISL_3050322 | EPI_ISL_2777972 |
| EPI_ISL_2777726 | EPI_ISL_2777811 | EPI_ISL_2777938 | EPI_ISL_3050551 | EPI_ISL_2777523 |
| EPI_ISL_2777727 | EPI_ISL_2777812 | EPI_ISL_2777939 | EPI_ISL_3050381 | EPI_ISL_2777531 |
| EPI_ISL_2777728 | EPI_ISL_2777815 | EPI_ISL_2777940 | EPI_ISL_3050415 | EPI_ISL_2777548 |
| EPI_ISL_2777729 | EPI_ISL_2777816 | EPI_ISL_2777948 | EPI_ISL_3050431 | EPI_ISL_2777973 |
| EPI_ISL_2777334 | EPI_ISL_2777817 | EPI_ISL_2777950 | EPI_ISL_3050529 | EPI_ISL_2777974 |
| EPI_ISL_2777730 | EPI_ISL_2777818 | EPI_ISL_2777295 | EPI_ISL_4030368 | EPI_ISL_2777975 |
| EPI_ISL_2777731 | EPI_ISL_2777819 | EPI_ISL_2777345 | EPI_ISL_4030369 | EPI_ISL_2777976 |
| EPI_ISL_2777732 | EPI_ISL_2777820 | EPI_ISL_2777952 | EPI_ISL_4030370 | EPI_ISL_2777568 |
| EPI_ISL_2777733 | EPI_ISL_2777821 | EPI_ISL_2777954 | EPI_ISL_4030371 | EPI_ISL_2777578 |
| EPI_ISL_2777734 | EPI_ISL_2777823 | EPI_ISL_2777955 | EPI_ISL_4030372 | EPI_ISL_2777579 |
| EPI_ISL_2777735 | EPI_ISL_2777827 | EPI_ISL_2777962 | EPI_ISL_4030373 | EPI_ISL_2777287 |
| EPI_ISL_2777736 | EPI_ISL_2777828 | EPI_ISL_2777964 | EPI_ISL_4030374 | EPI_ISL_2777582 |
| EPI_ISL_2777737 | EPI_ISL_2777829 | EPI_ISL_2777965 | EPI_ISL_4030375 | EPI_ISL_2777583 |
| EPI_ISL_2777738 | EPI_ISL_2777830 | EPI_ISL_2777243 | EPI_ISL_4030376 | EPI_ISL_2777587 |
| EPI_ISL_2777739 | EPI_ISL_2777833 | EPI_ISL_2777244 | EPI_ISL_4030377 | EPI_ISL_2777590 |
| EPI_ISL_2777740 | EPI_ISL_2777834 | EPI_ISL_2777245 | EPI_ISL_4030378 | EPI_ISL_2777597 |
| EPI_ISL_2777742 | EPI_ISL_2777837 | EPI_ISL_2777297 | EPI_ISL_4030379 | EPI_ISL_2777602 |
| EPI_ISL_2777743 | EPI_ISL_2777838 | EPI_ISL_2777318 | EPI_ISL_4030380 | EPI_ISL_2777608 |
| EPI_ISL_2777744 | EPI_ISL_2777839 | EPI_ISL_2777346 | EPI_ISL_4030381 | EPI_ISL_2777611 |
| EPI_ISL_2777745 | EPI_ISL_2777840 | EPI_ISL_2777347 | EPI_ISL_4030382 | EPI_ISL_2777977 |
| EPI_ISL_2777746 | EPI_ISL_2777841 | EPI_ISL_2777348 | EPI_ISL_4030324 | EPI_ISL_2777614 |
| EPI_ISL_2777747 | EPI_ISL_2777842 | EPI_ISL_4030383 | EPI_ISL_4030325 | EPI_ISL_2777978 |
| EPI_ISL_2777748 | EPI_ISL_2777843 | EPI_ISL_4030319 | EPI_ISL_4030326 | EPI_ISL_2777617 |
| EPI_ISL_2777749 | EPI_ISL_2777844 | EPI_ISL_3050307 | EPI_ISL_4030314 | EPI_ISL_2777625 |
| EPI_ISL_2777750 | EPI_ISL_2777845 | EPI_ISL_3050308 | EPI_ISL_4030327 | EPI_ISL_2777979 |

| | | | | |
|---|---|---|---|---|
| EPI_ISL_2777981 | EPI_ISL_3050351 | EPI_ISL_3050599 | EPI_ISL_3050528 | EPI_ISL_2777846 |
| EPI_ISL_2777982 | EPI_ISL_3050530 | EPI_ISL_3050342 | EPI_ISL_3050380 | EPI_ISL_2777853 |
| EPI_ISL_2777633 | EPI_ISL_3050531 | EPI_ISL_3050395 | EPI_ISL_3050438 | EPI_ISL_2777856 |
| EPI_ISL_2777983 | EPI_ISL_3050352 | EPI_ISL_3050606 | EPI_ISL_3050361 | EPI_ISL_2777860 |
| EPI_ISL_2777984 | EPI_ISL_3050323 | EPI_ISL_3050607 | EPI_ISL_3050362 | EPI_ISL_2777343 |
| EPI_ISL_2777640 | EPI_ISL_3050432 | EPI_ISL_3050608 | EPI_ISL_3050440 | EPI_ISL_2777869 |
| EPI_ISL_2777649 | EPI_ISL_3050575 | EPI_ISL_3050609 | EPI_ISL_3050452 | EPI_ISL_2777890 |
| EPI_ISL_2777652 | EPI_ISL_3050518 | EPI_ISL_3050423 | EPI_ISL_3050486 | EPI_ISL_2777893 |
| EPI_ISL_2777656 | EPI_ISL_3050642 | EPI_ISL_3050396 | EPI_ISL_3050487 | EPI_ISL_2777895 |
| EPI_ISL_2777660 | EPI_ISL_3050297 | EPI_ISL_3050397 | EPI_ISL_3050645 | EPI_ISL_2777901 |
| EPI_ISL_2777661 | EPI_ISL_3050353 | EPI_ISL_3050491 | EPI_ISL_3050646 | EPI_ISL_2777902 |
| EPI_ISL_2777929 | EPI_ISL_3050497 | EPI_ISL_3050390 | EPI_ISL_3050453 | EPI_ISL_2777906 |
| EPI_ISL_2777663 | EPI_ISL_3050505 | EPI_ISL_3050408 | EPI_ISL_3050454 | EPI_ISL_2777907 |
| EPI_ISL_2777666 | EPI_ISL_3050584 | EPI_ISL_3050424 | EPI_ISL_3050455 | EPI_ISL_2777344 |
| EPI_ISL_2777667 | EPI_ISL_3050416 | EPI_ISL_3050533 | EPI_ISL_3050456 | EPI_ISL_2777912 |
| EPI_ISL_2777671 | EPI_ISL_3050418 | EPI_ISL_3050469 | EPI_ISL_3050495 | EPI_ISL_2777918 |
| EPI_ISL_2777673 | EPI_ISL_3050402 | EPI_ISL_3050498 | EPI_ISL_3050496 | EPI_ISL_2777920 |
| EPI_ISL_2777674 | EPI_ISL_3050545 | EPI_ISL_3050493 | EPI_ISL_3050632 | EPI_ISL_2777921 |
| EPI_ISL_2777679 | EPI_ISL_3050638 | EPI_ISL_3050494 | EPI_ISL_3050633 | EPI_ISL_2777926 |
| EPI_ISL_2777316 | EPI_ISL_3050521 | EPI_ISL_3050536 | EPI_ISL_3050634 | EPI_ISL_2777927 |
| EPI_ISL_2777680 | EPI_ISL_3050441 | EPI_ISL_3050537 | EPI_ISL_3050649 | EPI_ISL_2777930 |
| EPI_ISL_2777681 | EPI_ISL_3050592 | EPI_ISL_3050523 | EPI_ISL_3050514 | EPI_ISL_2777933 |
| EPI_ISL_2777682 | EPI_ISL_3050442 | EPI_ISL_3050524 | EPI_ISL_3050543 | EPI_ISL_2777945 |
| EPI_ISL_2777685 | EPI_ISL_3050435 | EPI_ISL_3050525 | EPI_ISL_3050544 | EPI_ISL_2777949 |
| EPI_ISL_2777695 | EPI_ISL_3050554 | EPI_ISL_3050509 | EPI_ISL_3050552 | EPI_ISL_2777951 |
| EPI_ISL_2777359 | EPI_ISL_3050459 | EPI_ISL_3050527 | EPI_ISL_2778008 | EPI_ISL_2777953 |
| EPI_ISL_2777332 | EPI_ISL_3050460 | EPI_ISL_3050328 | EPI_ISL_2778009 | EPI_ISL_2777957 |
| EPI_ISL_2777753 | EPI_ISL_3050461 | EPI_ISL_3050588 | EPI_ISL_3050386 | EPI_ISL_2777960 |
| EPI_ISL_2777756 | EPI_ISL_3050580 | EPI_ISL_3050589 | EPI_ISL_3050546 | EPI_ISL_2777288 |
| EPI_ISL_2777760 | EPI_ISL_3050581 | EPI_ISL_3050590 | EPI_ISL_2777442 | EPI_ISL_2777349 |
| EPI_ISL_2777773 | EPI_ISL_3050316 | EPI_ISL_3050383 | EPI_ISL_2777447 | EPI_ISL_2777350 |
| EPI_ISL_2777774 | EPI_ISL_3050547 | EPI_ISL_3050569 | EPI_ISL_2777514 | EPI_ISL_2777351 |
| EPI_ISL_2777778 | EPI_ISL_3050317 | EPI_ISL_3050600 | EPI_ISL_2777529 | EPI_ISL_2777352 |
| EPI_ISL_2777810 | EPI_ISL_3050331 | EPI_ISL_3050602 | EPI_ISL_2777550 | EPI_ISL_2777353 |
| EPI_ISL_2777813 | EPI_ISL_3050557 | EPI_ISL_3050472 | EPI_ISL_2777555 | EPI_ISL_2777354 |
| EPI_ISL_2777814 | EPI_ISL_3050559 | EPI_ISL_3050611 | EPI_ISL_2777577 | EPI_ISL_2777355 |
| EPI_ISL_2777824 | EPI_ISL_3050560 | EPI_ISL_3050500 | EPI_ISL_2777580 | EPI_ISL_2777968 |
| EPI_ISL_2777825 | EPI_ISL_3050356 | EPI_ISL_3050612 | EPI_ISL_2777592 | EPI_ISL_3050616 |
| EPI_ISL_2777852 | EPI_ISL_3050346 | EPI_ISL_3050603 | EPI_ISL_2777594 | EPI_ISL_3050574 |
| EPI_ISL_2777854 | EPI_ISL_3050329 | EPI_ISL_3050625 | EPI_ISL_2777596 | EPI_ISL_3050458 |
| EPI_ISL_2777865 | EPI_ISL_3050365 | EPI_ISL_3050626 | EPI_ISL_2777598 | EPI_ISL_3050488 |
| EPI_ISL_2777866 | EPI_ISL_3050388 | EPI_ISL_3050628 | EPI_ISL_2777607 | EPI_ISL_3050504 |
| EPI_ISL_2777887 | EPI_ISL_3050358 | EPI_ISL_3050629 | EPI_ISL_2777610 | EPI_ISL_3050572 |
| EPI_ISL_2777891 | EPI_ISL_3050404 | EPI_ISL_3050637 | EPI_ISL_2777613 | EPI_ISL_3050573 |
| EPI_ISL_2777904 | EPI_ISL_3050338 | EPI_ISL_3050571 | EPI_ISL_2777619 | EPI_ISL_3050576 |
| EPI_ISL_2777905 | EPI_ISL_3050420 | EPI_ISL_3050613 | EPI_ISL_2777623 | EPI_ISL_3050577 |
| EPI_ISL_2777908 | EPI_ISL_3050586 | EPI_ISL_3050296 | EPI_ISL_2777629 | EPI_ISL_3050578 |
| EPI_ISL_2777909 | EPI_ISL_3050587 | EPI_ISL_3050614 | EPI_ISL_2777644 | EPI_ISL_3050579 |
| EPI_ISL_2777910 | EPI_ISL_3050359 | EPI_ISL_3050368 | EPI_ISL_2777648 | EPI_ISL_3050553 |
| EPI_ISL_2777914 | EPI_ISL_3050489 | EPI_ISL_3050369 | EPI_ISL_2777653 | EPI_ISL_3050382 |
| EPI_ISL_2777924 | EPI_ISL_3050462 | EPI_ISL_3050370 | EPI_ISL_2777664 | EPI_ISL_3050364 |
| EPI_ISL_2777925 | EPI_ISL_3050463 | EPI_ISL_3050371 | EPI_ISL_2777665 | EPI_ISL_3050433 |
| EPI_ISL_2777928 | EPI_ISL_3050467 | EPI_ISL_3050385 | EPI_ISL_2777672 | EPI_ISL_3050417 |
| EPI_ISL_2777931 | EPI_ISL_3050318 | EPI_ISL_3050392 | EPI_ISL_2777677 | EPI_ISL_3050520 |
| EPI_ISL_2777941 | EPI_ISL_3050334 | EPI_ISL_3050640 | EPI_ISL_2777684 | EPI_ISL_3050443 |
| EPI_ISL_2777942 | EPI_ISL_3050325 | EPI_ISL_3050393 | EPI_ISL_2777688 | EPI_ISL_3050355 |
| EPI_ISL_2777943 | EPI_ISL_3050561 | EPI_ISL_3050474 | EPI_ISL_2777694 | EPI_ISL_3050555 |
| EPI_ISL_2777944 | EPI_ISL_3050582 | EPI_ISL_3050475 | EPI_ISL_2777712 | EPI_ISL_3050585 |
| EPI_ISL_2777985 | EPI_ISL_3050421 | EPI_ISL_3050510 | EPI_ISL_2777329 | EPI_ISL_3050507 |
| EPI_ISL_2777946 | EPI_ISL_3050619 | EPI_ISL_3050511 | EPI_ISL_2777335 | EPI_ISL_3050556 |
| EPI_ISL_2777956 | EPI_ISL_3050620 | EPI_ISL_3050630 | EPI_ISL_2777336 | EPI_ISL_3050567 |
| EPI_ISL_2777958 | EPI_ISL_3050300 | EPI_ISL_3050631 | EPI_ISL_2777337 | EPI_ISL_3050532 |
| EPI_ISL_2777959 | EPI_ISL_3050621 | EPI_ISL_3050409 | EPI_ISL_2777338 | EPI_ISL_3050548 |
| EPI_ISL_2777991 | EPI_ISL_3050622 | EPI_ISL_3050540 | EPI_ISL_2777339 | EPI_ISL_3050324 |
| EPI_ISL_2777961 | EPI_ISL_3050604 | EPI_ISL_3050349 | EPI_ISL_2777761 | EPI_ISL_3050549 |
| EPI_ISL_2777966 | EPI_ISL_3050639 | EPI_ISL_3050379 | EPI_ISL_2777770 | EPI_ISL_3050345 |
| EPI_ISL_2777967 | EPI_ISL_3050305 | EPI_ISL_3050437 | EPI_ISL_2777341 | EPI_ISL_3050405 |
| EPI_ISL_3050457 | EPI_ISL_3050326 | EPI_ISL_3050478 | EPI_ISL_2777342 | EPI_ISL_3050419 |
| EPI_ISL_3050515 | EPI_ISL_3050311 | EPI_ISL_3050480 | EPI_ISL_2777777 | EPI_ISL_3050295 |
| EPI_ISL_3050591 | EPI_ISL_3050327 | EPI_ISL_3050481 | EPI_ISL_2777809 | EPI_ISL_3050304 |
| EPI_ISL_3050641 | EPI_ISL_3050594 | EPI_ISL_3050482 | EPI_ISL_2777822 | EPI_ISL_3050465 |
| EPI_ISL_3050615 | EPI_ISL_3050595 | EPI_ISL_3050483 | EPI_ISL_2777826 | EPI_ISL_3050466 |
| EPI_ISL_3050516 | EPI_ISL_3050522 | EPI_ISL_3050484 | EPI_ISL_2777831 | EPI_ISL_3050508 |
| EPI_ISL_3050517 | EPI_ISL_3050468 | EPI_ISL_3050485 | EPI_ISL_2777832 | EPI_ISL_3050299 |
| EPI_ISL_3050617 | EPI_ISL_3050341 | EPI_ISL_3050643 | EPI_ISL_2777835 | EPI_ISL_3050339 |
| EPI_ISL_3050618 | EPI_ISL_3050597 | EPI_ISL_3050644 | EPI_ISL_2777836 | EPI_ISL_3050310 |

EPI_ISL_3050340
EPI_ISL_3050306
EPI_ISL_3050333
EPI_ISL_3050596
EPI_ISL_3050320
EPI_ISL_3050598
EPI_ISL_3050366
EPI_ISL_3050389
EPI_ISL_3050347
EPI_ISL_3050610
EPI_ISL_3050422
EPI_ISL_3050492
EPI_ISL_3050534
EPI_ISL_3050535
EPI_ISL_3050526
EPI_ISL_3050550
EPI_ISL_3050568
EPI_ISL_3050623
EPI_ISL_3050473
EPI_ISL_3050539
EPI_ISL_3050348
EPI_ISL_3050372
EPI_ISL_3050313
EPI_ISL_3050321
EPI_ISL_3050343
EPI_ISL_3050330
EPI_ISL_3050335
EPI_ISL_3050336
EPI_ISL_3050360
EPI_ISL_3050373
EPI_ISL_3050374
EPI_ISL_3050377
EPI_ISL_3050378
EPI_ISL_3050398
EPI_ISL_3050399
EPI_ISL_3050400
EPI_ISL_3050410
EPI_ISL_3050411
EPI_ISL_3050412
EPI_ISL_3050413
EPI_ISL_3050425
EPI_ISL_3050426
EPI_ISL_3050427
EPI_ISL_3050428
EPI_ISL_3050429
EPI_ISL_3050430
EPI_ISL_3050414
EPI_ISL_3050444
EPI_ISL_3050445
EPI_ISL_3050446
EPI_ISL_3050447
EPI_ISL_3050476
EPI_ISL_3050477
EPI_ISL_3050512
EPI_ISL_3050513
EPI_ISL_3050448
EPI_ISL_3050449
EPI_ISL_3050450
EPI_ISL_3050451
EPI_ISL_3050541
EPI_ISL_3050542
EPI_ISL_3050583
EPI_ISL_3050562
EPI_ISL_3050479
EPI_ISL_3050439
EPI_ISL_3050563
EPI_ISL_3050635
EPI_ISL_3050647
EPI_ISL_3050648
EPI_ISL_3050650
EPI_ISL_3050314
EPI_ISL_3050564
EPI_ISL_3050350
